# Representing multimorbid disease progressions using directed hypergraphs

**DOI:** 10.1101/2023.08.31.23294903

**Authors:** Jamie Burke, Ashley Akbari, Rowena Bailey, Kevin Fasusi, Ronan A. Lyons, Jonathan Pearson, James Rafferty, Daniel Schofield

## Abstract

**Objective:** To introduce directed hypergraphs as a novel tool for assessing the temporal relationships between coincident diseases, addressing the need for a more accurate representation of multimorbidity and leveraging the growing availability of electronic healthcare databases and improved computational resources.

**Methods:** Directed hypergraphs offer a high-order analytical framework that goes beyond the limitations of directed graphs in representing complex relationships such as multimorbidity. We apply this approach to multimorbid disease progressions observed from two multimorbidity sub-cohorts of the SAIL Databank, after having been filtered according to the Charlson and Elixhauser comorbidity indices, respectively. After constructing a novel weighting scheme based on disease prevalence, we demonstrate the power of these higher-order models through the use of PageRank centrality to detect and classify the temporal nature of conditions within the two comorbidity indices.

**Results:** In the Charlson population, we found that chronic pulmonary disease (CPD), cancer and diabetes were conditions observed early in a patient’s disease progression (predecessors), with stroke and dementia appearing later on (successors) and myocardial infarction acting as a transitive condition to renal failure and congestive heart failure. In Elixhauser, we found renal failure, neurological disorders and arrhythmia were classed as successors and hypertension, depression, CPD and cancer as predecessors, with diabetes becoming a transitive condition in the presence of obesity and alcohol abuse. The dynamics of these and other conditions changed across age and sex but not across deprivation. Unlike the directed graph, the directed hypergraph could model higher-order disease relationships, which translated into stronger classifications between successor and predecessor conditions, alongside the removal of spurious results.

**Conclusion:** This study underscores the utility of directed hypergraphs as a powerful approach to investigate and assess temporal relationships among coincident diseases. By overcoming the limitations of traditional pairwise models, directed hypergraphs provide a more accurate representation of multimorbidity, offering insights that can significantly contribute to healthcare decision-making, resource allocation, and patient management. Further research holds promise for advancing our understanding of critical issues surrounding multimorbidity and its implications for healthcare systems.

## 1 Introduction

Multimorbidity is the health state of having two or more concurrent conditions [1]. This is becoming more common as populations age through increased survival rates from acute and chronic illness due to advancements in technology and healthcare. However, multimorbidity is poorly understood and is combated by a compartmentalised healthcare system, originally designed under a single-disease framework. This model leads to fragmented, costly and ineffective care, leading to individuals living with multimorbidity not getting the complex care they need [2, 3]. Improving our understanding of multimorbidity may help inform clinical decisions to improve patient services and outcomes, as well as help relieve some of the burden on healthcare services and available resources.

The collection of anonymised individual-level, population-scale, linked electronic health record (EHR) data sources available within a single trusted research environment (TRE) creates several opportunities for insight and analysis into multimorbidity investigation. The Secure Anonymised Information Linkage (SAIL) Databank provides access to anonymised individual-level, population-scale, linked EHR data sources for the purpose of research, powered by its secure e-research platform (SERP) [4, 5, 6]. Derived from SAIL was the Wales Multimorbidity e-Cohort (WMC) [7], created to provide an accessible, research ready data asset (RRDA) to further the understanding of multimorbidity.

With access to population-scale EHR data, and as the pattern of health and disease is changing in our population [8], there is a growing focus on establishing statistical and analytical frameworks to sufficiently model population-level multimorbidity cohorts. Greater availability of computational resources and a diverse set of analytical approaches has ushered in a range of analyses on multimorbidity data, which includes disease and population clustering [9, 10, 11, 12, 13, 14] and network medicine [15, 16, 17, 18, 19].

However, the majority of these studies take a cross-sectional perspective to investigating multimorbidity. While snap-shot analyses are helpful, modelling disease and multimorbidity trajectories better represent the development of multimorbidity in populations and could have stronger implications on policy making and strategy decisions within healthcare services. Indeed, approaches to visualise, identify patterns from, and model longitudinal multimorbidity data are emerging, and a recent review [20] identified 35 such studies focusing on analytical approaches to longitudinal multimorbidity data.

Of these 35 publications identified, [20] highlighted 9 academic papers on transition and disease progression modelling. These publications included simple temporal correlation analysis [21, 22], Markov chain and multi-state modelling [23, 24, 25, 26], multi-level temporal Bayesian networks [27, 28] and network analysis [17, 18, 29]. Siah, et al. [17, 18] performed a comprehensive analysis of UK multimorbidity data with directed simple graphs and using centrality to analyse different stratification’s of their data using different demographic factors. This work is similar to our study but limited by only being able to assess pairwise, comorbid relationships between diseases.

A recent paper [30] established a hypergraph framework allowing the application of higher complexity to multimorbidity data. Rafferty, et al. investigated the use of multimorbidity prevalence and graph centrality to identify sets of chronic diseases which have an inordinate effect on population prevalence [30]. Hypergraphs are ideally suited as a modelling tool for multimorbidity, acting as a natural extension to simple graphs modelling comorbidity. Our study sought to extend upon this work by introducing directionality into the hypergraph to model multimorbidity disease progression and assess the contributory relationships between coincident diseases. While directed simple graphs allow for modelling comorbidity disease progressions [17, 18], directed hypergraphs allow for the modelling of multimorbidity disease progressions.

A key benefit of directed hypergraphs is that they can be built without any requirement of temporality or probabilities, and can be analysed using known techniques in the field of graph theory. In comparison to the directed graph, Markov chain and multi-state models, directed hypergraphs provide a greater degree of flexibility in its edge weight system as well as its diverse range of downstream tasks on the structure itself. In particular, the directed graph is a subgraph of the directed hypergraph, the Markov chain model can be generated from the directed hypergraph through the inclusion of probabilistic edges or using random walk theory, and the inclusion of temporality and demographic factors can be used to restructure the graph into a multi-state model. From here, a range of diverse analyses can be performed, including survival modelling or assessment of population-specific risk factors linked to multimorbidity development. Multilevel Bayesian networks model hierarchical, structured patient care data, grouping demographic factors separate from diseases for conditional dependence and disease prediction [31, 28]. We believe this approach could benefit from using directed hypergraphs to model temporality and prevalence among multimorbidity development, including calculation of transition rates between exact diseases rather than the number of morbidities observed [28].

We demonstrate the use case for directed hypergraphs by assessing the contributory relationships chronic health conditions have with one another through the lens of multimorbidity prevalence. Through PageRank, a centrality-based metric, we can assign directional relationships between diseases through observed patient disease progressions. We stratify our data to understand these relationships at different population subgroup levels using demographic variables such as age, sex and deprivation status, three major contributors to multimorbidity [32]. This study provides insight into a powerful and flexible model for analysing longitudinal multimorbidity data, which could have important implications on policy and strategy decisions within healthcare services.

**Problem** Multimorbidity and longitudinal multimorbidity are often poorly represented by methodologies which cannot accurately represent the complex, multi-disease relationships that patients experience.

**What is Already Known** Hypergraphs have recently been used to model multimorbidity in the undirected case, and are presented as a better model for disease interaction in multimorbidity over simple graphs. Directed hypergraphs however have never been applied to model longitudinal multimorbidity data.

**What This Paper Adds** We introduce the directed hypergraph architecture into the field of multimorbidity research. We demonstrate the power of directed hypergraphs to overcome the limitations of directed graph equivalents and provide an open-source model implementation to facilitate further collaboration.

### 1.1 Background

In general, graphs model the relationships between objects, called nodes, through pairwise connections, called edges. In the context of multimorbidity, the nodes commonly represent diseases, and the edges represent co-occurring pairs of diseases or comorbidities [15, 16, 19]. The simple graph only allows for two nodes to be connected using at most one edge, and these edges are typically weighted according to some outcome of interest, such as disease prevalence. A directed graph imposes directionality among the edges. Here, nodes can have incoming and outgoing connections and edge weights are related to its direction. We can relate the undirected graph to the directed graph by noting that each undirected parent edge gives rise to at most two children directed edges^1^. In multimorbidity, a directed graph represents co-morbid disease progressions [17, 18]. The relationship between the undirected and directed hypergraph are similar, but now permit any number of nodes to be connected through a single edge, acting as a more natural modelling tool in the context of multimorbidity. See section 10.1 in the supplementary materials for an example to accompany this background section using synthetic data.

#### 1.1.1 Hypergraphs and directed hypergraphs

Let ℋ(*V, E, W_E_, W_V_*) define an undirected hypergraph with node set *V* = {*v*_1_*, v*_2_*, …, v_n_*} and hyperedge set *E* = {*e*_1_*, e*_2_*, …, e_m_*}, with *n* = |*V* | nodes and *m* = |*E*| hyperedges. The hyperedges are arbitrary subsets of *V*, with weight *W* (*e_i_*) associated with hyperedge *e_i_*. Hyperedge weights are collected along the diagonal of an *m* × *m* diagonal matrix *W_E_*. Similarly, *W_V_* is an *n* × *n* diagonal matrix storing the node weights. In the subsequent analyses performed, we do not make use of the node weight matrix, and will not be mentioned henceforth. A fundamental representation of the hypergraph is the *n* × *m* incidence matrix *M* which stores information on which nodes each hyperedge connects. For some node *v_i_* and hyperedge *e_j_*, *M* (*v_i_, e_j_*) = 1 if *v_i_* and *e_j_* are incident. We refer to Rafferty, et al. [30] for a more detailed description on undirected hypergraphs.

A directed hypergraph ℋ*_D_*(*V,* ε *, W_ε_*) is a collection of the same node set *V* as above, a set of *directed* hyperedges, or *hyperarcs*, ε = {*h*_1_*, …, h_k_*}. Each hyperarc *h_i_* = ⟨*T* (*h_i_*)*, H*(*h_i_*)⟩ where *T* (*h_i_*)*, H*(*h_i_*) ⊆ *V* represents a collection of nodes such that those *v_i_*∈ *T* (*h_i_*) are the tails of the hyperarc and those *v_j_* ∈ *H*(*h_i_*) are the heads of the hyperarc. The tail set of *h_i_*are nodes with outgoing connections and the head set are nodes with incoming connections of *h_i_*. Each hyperarc *h_i_*is associated with a weight *w*(*h_i_*), stored in the diagonal of a *k* × *k* diagonal matrix *W_E_* .

In multimorbidity, *V* = {*D*_1_*, …, D_n_*} represents the set of chronic health conditions being modelled. In the undirected hypergraph ℋ, hyperedges *e_i_* ∈ *E* represent multimorbidity disease sets while in the directed hypergraph H*_D_*, hyperarcs *h_i_* ∈ E represent multimorbidity disease progression. We assume that multimorbidity disease progression is an incremental, irreversible process. In this sense, a participant’s diseases are never “cured,” and they can only acquire more diseases, aggregating individual conditions to their disease trajectory according to their observed disease progression. By this logic, we only consider hyperarcs which have a single incoming connection (also known as B-hyperarcs), such that |*H*(*h_i_*)| = 1 for *i* = 1*, …, k*. For example, given a multimorbidity disease set {*D*_1_*, D*_2_*, D*_3_}, whose ordering of observed diagnoses follows *D*_1_ → *D*_2_ → *D*_3_, we construct 2 hyperarcs to represent this multimorbidity disease progression, *h*_1_ = ⟨{*D*_1_}, {*D*_2_}⟩ representing *D*_1_ → *D*_2_, and *h*_2_ = ⟨{*D*_1_*, D*_2_}, {*D*_3_}⟩ representing *D*_1_*, D*_2_ → *D*_3_.

Figure 1 shows four different unweighted, fully connected graphs with three disease nodes. Note that the undirected and directed hyperedges in magenta seen in figures 1B and 1D are not possible to model using the simpler graph models in figures 1A and 1C. Comparing the undirected graphs with their directed equivalent, the undirected edges in figures 1A and 1B give rise to multiple directed edges in figures 1C and 1D. This is particularly important when considering weighted graph models. We can also observe the existence of self-looping directed edges in 1C and 1D. In the context of multimorbidity, these represent individuals with only a single condition observed throughout the period of analysis.

**Figure 1:**
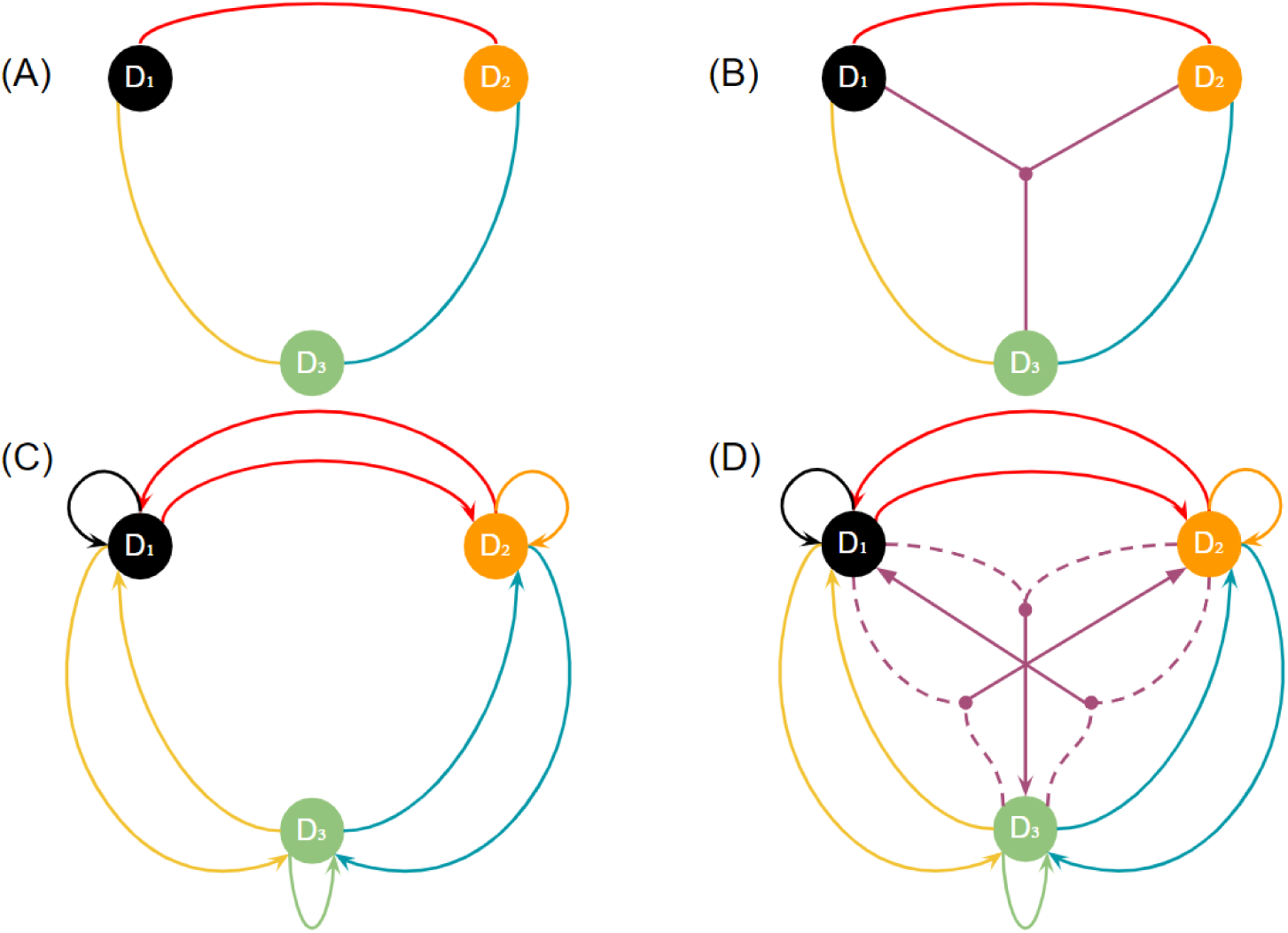
Four different types of unweighted, fully connected graph models with 3 nodes. (A) undirected graph, (B) undirected hypergraph, (C) directed graph, (D) directed hypergraph of B-hyperarcs, the only type of hyperarc considered in this work. Dotted lines here represent nodes as part of the tail set in each hyperarc. Edges have been colour coded to help identify children from their parents — looking top-down, we can observe how the directed edges (children) are generated from their corresponding undirected edges (parents). Note also the existence of self-edges in the directed representations.

#### 1.1.2 Hyperedge and hyperarc weighting

Hyperarcs are generated directly from their parent, undirected hyperedge. In particular, given an undirected (parent) hyperedge {*D*_1_*, D*_2_*, D*_3_} with three nodes, we can define three possible hyperarcs (children) based on this hyperedge, namely: *D*_1_*, D*_2_ → *D*_3_; *D*_2_*, D*_3_ → *D*_1_; and *D*_1_*, D*_3_ → *D*_2_. Because of this directed relationship between hyperedges and hyperarcs, we first outline the rationale for computing hyperedge weights for an undirected hypergraph. Each hyperarc weight is then computed directly from their parent.

The choice of the edge weights in a graph determines the interpretation of the analysis. We use disease prevalence as the outcome of interest when quantifying the relationship coincident diseases have with each other. Multimorbidity prevalence is an important metric to quantify and track longitudinally. Quantitative insights on such population-level statistics can help inform which disease sets and progressions are having a disproportionate impact on population subgroups.

In network medicine, it is common to use metrics such as odds ratio, lift [15, 17, 18] or partial Pearson correlation [33, 19] to measure pairwise connectivity from comorbid disease prevalence. However, these and other common metrics for simple graphs cannot be applied when measuring the similarity of multiple groups. In multimorbidity, the hyperedge weight measures the relative prevalence of a disease set and is some normalised measure of overlapping sets. We adopt an extension to the Sørensen-Dice similarity coefficient [34, 35] to measure the prevalence of a multimorbidity disease set, relative to other multimorbidity disease sets.

Let *C*(*e_i_*) represent the raw prevalence count of disease set *e_i_*. This value represents the number of individuals who were observed to have all diseases in *e_i_*. The weight of hyperedge *e_i_*, *W* (*e_i_*), is the multi-set overlap

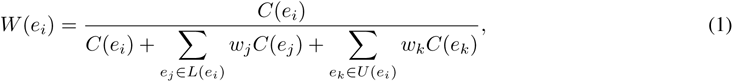

where

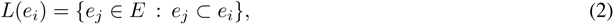

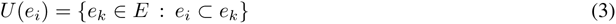

are all proper subsets and supersets of *e_i_* (related to the power and super-power set, respectively). In particular, *L*(*e_i_*) contains all disease sets which are a proper subset of the diseases in *e_i_* (with *C*(∅⊘) = 0 for the empty set). The set *U* (*e_i_*) contains all disease sets which already contain all diseases in *e_i_*, and other diseases. This formulation^2^ penalises an observed count of contributions from individuals for a disease set by all other disease sets which are either a subset or super set of the disease set in question. A hyperedge’s weight is penalised by the largest possible number of individuals who have some temporal link to that disease set and is a normalised and relative measure of how pervasive a disease set trajectory is among the population which have had similar disease set trajectories. While the weight coefficients in equation (1) are arbitrary, in our work we have set *w_j_* = *w_k_* = 1 for all *j, k*. This is so there is equal penalisation across all proper subsets and supersets of the disease set.

An individual will contribute to a hyperedge’s raw prevalence count, *C*(*e_i_*) if they had all diseases in *e_i_* during their multimorbidity disease progression. For example, given an individual’s multimorbidity progression *D*_1_ → *D*_2_ → *D*_3_, they would contribute a single unit of prevalence to the hyperedges in {*D*_1_}, {*D*_1_*, D*_2_}, {*D*_1_*, D*_2_*, D*_3_}. In the directed hypergraph case, they would contribute a single unit of prevalence to the hyperarcs in {{*D*_1_ → *D*_2_}, {*D*_1_*, D*_2_ → *D*_3_}}. For the hyperarc weighting system, there should be consideration of both the prevalence of the hyperarc itself, i.e. the progression it represents, as well as the parent hyperedge to which it is a child of. Let *P* (E) be the set of parent hyperedges to all hyperarcs in ε, i.e. *P*(ε) = *E* from the undirected hypergraph. Then, hyperarc *h_i_* has parent *p*(*h_i_*) ∈ *P*(ε) with K(*h_i_*) = {*h_j_* : *p*(*h_j_*) = *p*(*h_i_*)} as the set of siblings of hyperarc *h_i_*, i.e. all hyperarcs which share the same parent hyperedge. The weight for hyperarc *h_i_*, *w*(*h_i_*), is

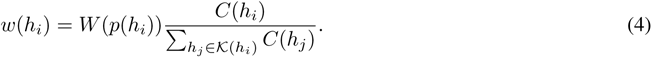

Note that *W* (·) represents a *hyperedge* weight in the undirected hypergraph model, while *w*(·) represents a *hyperarc* weight. Here, the hyperarc weighting measures how prevalent the hyperarc *h_i_*is among its siblings with the same parent relative to how prevalent that parent hyperedge is among the population. That is, it measures how prevalent an observed disease progression is among other disease progressions possible from the same multimorbidity set, relative to the overall prevalence of that multimorbidity set within the population.

#### 1.1.3 PageRank for directed hypergraphs

The directed nature of the connections in directed hypergraphs give us the ability to measure the degree of incoming and outgoing connectivity each disease node has, i.e., how many times it appears as the oncoming diagnosis (head node), or a previous diagnosis (tail node) in a disease progression (hyperarc). In multimorbidity, understanding the temporal nature of each disease node based on their connectivity would be useful for understanding disease progression and any contributory relationships between diseases.

PageRank centrality in directed graphs measures node popularity based on the number of incoming connections that each node has, the weight of those incoming connections, and the weight of the incoming connections connected to those aforementioned connections. PageRank was originally developed for the World Wide Web to determine the importance of webpages related to a given query, based on the incoming connectivity of other webpages (via hyperlinks), and the connectivity of those aforementioned webpages on the World Wide Web [36].

PageRank in directed graphs more generally measures the transitive influence of each nodes’ incoming connectivity. In brief, PageRank computes the principal left eigenvector of the directed hypergraphs’ probability transition matrix (PTM). This PTM measures the transition rates between pairwise nodes in the network based on random walk theory [37]. The dominant eigenvector of this PTM, called the PageRank vector, computes the long-term probability of reaching a target node, independent of the source node. Since the PTM is stochastic, the principal left eigenvector has all positive entries by the Perron-Frobenium theorem [38], and all entries sum to 1 inducing a probability space. We can use this probability space to rank nodes in order of how likely the node is to be visited or reached in the network.

The PageRank algorithm has been described and developed for the directed hypergraph [39] and requires computing the PTM of the directed hypergraph. Using random walk theory, the PTM computes transition rates following the direction of the hyperarc, and it follows that PageRank measures node influence based on *incoming* connectivity. In multimorbidity, this represents the probability of diagnosis for a particular disease, given some previous morbidity. The PageRank vector computes a ranking of probabilities of how likely each disease in the network acts as a *successor* morbidity to other chronic health conditions. This successor-PageRank vector can be used for detecting important successor diseases, i.e. those diseases which are diagnosed subsequently to other conditions. The disease with the highest probability in the PageRank vector is the one which most frequently appears in the data as a subsequent diagnosis. Ducournau and Bretto [40] defined this successor PTM, *P_s_*for the directed hypergraph as

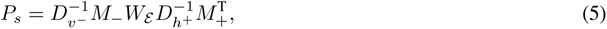

where *D_v−_* and *D_h_*_+_ are the tail and head degree matrices of the nodes and hyperarc, respectively and *M_−_* represents the incidence matrix for the tails and heads of all hyperarcs^3^. Note however that we only consider B-hyperarcs where the head degree of any hyperarc *h* is 1. Therefore, *D_h_*_+_ = I*_k_* where I*_k_* is the *k* × *k* identity matrix.

We also propose a simple formula rearrangement to permit measuring node influence based on *outgoing* connectivity. Given a current position *u* ∈ *V* in the directed hypergraph, we choose a hyperarc *h* ∈ E such that *u* ∈ *H*(*h*) with a probability proportional to *w*(*h*). Following random walk theory [37], a transition can only be made following the inverse direction of a hyperarc, i.e. from the head node to any member of its tail. We therefore choose a node *v* ∈ *T* (*h*) uniformly at random with probability proportion to 1*/δ_−_*_(_*_h_*_)_. The matrix notation for this PTM *P_p_*is defined by

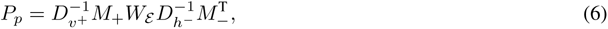

where *D_v_*_+_ and *D_h−_* represent the node head degree and hyperarc tail degree matrices of the undirected hypergraph [30]. In matrix form the PTM *P_p_* is a row-stochastic matrix and the proof is identical to the one by Tran, et al. [39]. According to this transition rule, we would interpret a transition probability *P_p_*(*i, j*) to be the probability that disease *j* was diagnosed prior to disease *i*. Computing the PageRank vector here would help us detect central disease nodes which act as important predecessor conditions for other diseases. The disease with the highest probability in the PageRank vector will represent health conditions at the start of a patient’s multimorbidity development, flagged by the healthcare system prior to many other subsequent diagnoses.

## 2 Material and methods

### 2.1 Data

The data and subsequent analyses was made possible through the SAIL Databank, a privacy-protecting TRE (an example of a Secure Data Environment (SDE)) [5, 6], providing secure access to a rich collection of de-identified anonymised primary and secondary care EHR data sources relating to the Welsh population. The SAIL Databank is one of the most comprehensive data sources in the UK, providing linkage at the individual and household level [4] through multiple EHR data sources, including critical care, general practice and inpatient data.

The dataset accessed and analysed from the SAIL Databank used in this study was the Welsh multimorbidity e-cohort (WMC) [7]. The WMC was created and derived from multiple demographic, administrative and EHR data sources relating to 2,902,101 individuals resident and receiving services in Wales. From all data sources, the total follow-up coverage is from the 1st January 2000 until either the 31st December 2019, a Welsh residency break or death, whichever was earliest. The primary and secondary care data sources also provide rich historical records to track disease progression over the course of an individuals lifetime, including historical EHR’s prior to 2000. This large multimorbidity cohort supersedes sample size, demographic and follow-up coverage compared to almost all identified studies in [20]. This study was performed under Project 1392 - Transforming healthcare data with graph-based techniques using the SAIL Databank; reviewed and approved by the Information Governance Review Panel (IGRP) of the SAIL Databank and Swansea University.

### 2.2 Variables

Among other code lists and processed data tables, two published comorbidity indices were used to measure and conceptualise multimorbidity in the WMC e-cohort using Read codes (for primary care) and International Classification of Diseases version 10 (ICD-10) codes (for secondary care) and were grouped into 16 and 30 condition categories, known as the Charlson [41, 42] and Elixhauser [43, 42] comorbidity indices, respectively — two popular and standardised comorbidity indices [44]. Their disease definitions have been used to classify read and ICD-10 codes into higher-level disease categories to build two distinct data tables, flagging date-time instances where individuals were first observed to have one or more of these conditions found in either index during an interaction with the healthcare system in Wales. Supplementary tables S2 and S4 show the comorbidities from each comorbidity index and can be found in the appendix.

The 16 conditions in the Charlson comorbidity table were reduced to 13 to prevent pseudo-clustering in the results. The three related sets of diseases were (i) cancer, lymphoma and leukaemia and metastatic cancer, (ii) mild and severe liver disease and (iii) diabetes with and without complication. Similarly, the 30 conditions in the Elixhauser disease table were reduced to 27 comorbidities. The three sets of diseases were (i) hypertension with and without complication, (ii) diabetes with and without complication and (iii) lymphoma and metastatic cancer. HIV/AIDS status is omitted from both comorbidity tables in the WMC to conform with SAIL policies as it was considered sensitive information by Wales in 2008 [7].

Our analysis is two-fold, using both comorbidity disease tables as the set of nodes in each directed hypergraph model. The latter, Elixhauser comorbidity table includes 30 diseases which is more than 80% of studies identified in [20]. We use the terms Charlson and Elixhauser populations to mean the population tables generated from the WMC, using either the Charlson comorbidity index or Elixhauser comorbidity index, respectively.

### 2.3 Participants

Participants from the WMC were included in the analysis if they were alive from January 1st 2000 and aged 20 years old or above according to the Medical Research Council (MRC) funded Welsh Multimorbidity Machine Learning (WMML) project [7]. The Charlson and Elixhauser populations were finalised by only studying subcohorts who all had at least one condition defined by the Charlson/Elixhauser comorbidity indexes. A diagnosis of a condition was made for an individual if there existed a date-time interaction from any WMC linked data source which indicated the incidence of said condition. We define the date-time instance to be the earliest interaction with the healthcare system which makes reference to this condition.

The tabular data of individuals and their date-time diagnoses are observations, rather than true realisations. Therefore, date-time values were used only to generate an observed ordering for each individual’s multimorbidity disease progression. This study did not use the specific date-time values to measure time between observed diagnoses. This is because longitudinal data is rarely complete, with follow-up visits scheduled intermittently and exact time of disease onset cannot be recorded explicitly.

Using the rule to set date-time instances for observed disease diagnoses, most individuals have a distinct multimorbidity progression, where observed diagnoses were made on unique dates. Their progression set and consequential contribution to the prevalence count of multimorbidity disease progression is clear. However, it is possible for an individual to have been diagnosed with multiple conditions on the same day.

This can be resolved by constructing their progression sets by considering all permuted trajectories of those duplicate conditions. For example, given an individual’s multimorbidity progression (*D*_1_ ↔ *D*_2_) → *D*_3_, i.e. diseases *D*_1_ and *D*_2_ were diagnosed on the same day, then the directed progression set is { {*D*_1_ → *D*_2_*, D*_2_ → *D*_1_}, {*D*_1_*, D*_2_ → *D*_3_} }. To account for the duplicate, the individual contributes a fraction of unit prevalence proportional to the number of permutations required to explain all possible trajectories. In this case, the individual contributes 1/2 to *D*_1_ → *D*_2_ and *D*_2_ → *D*_1_ and 1 to *D*_1_*, D*_2_ → *D*_3_. Note that we ignore the observed ordering of conditions in the tail set of hyperarcs.

For individuals where only two conditions were diagnosed on the same date, the correction can be made through halving the individuals contribution of unit prevalence to the two permuted progressions. However, for individuals with three or more conditions diagnosed on the same date, the number of permutations rise, making the individual’s multimorbidity trajectory less clear as their effective prevalence contribution becomes less significant. Therefore, we excluded these individuals. This resulted in 14,618 (1.1%) and 69,645 (4.1%) individuals excluded from the Charlson and Elixhauser populations, respectively, representing small proportions of the overall populations. See supplementary figure S4 for a cohort specification flowchart on how the Charlson and Elixhauser populations were defined.

### 2.4 Statistical analysis

Building the directed hypergraph first required calculating the hyperedge and and hyperarc weights. This was accomplished by extracting the records for each individual participant, ordering their conditions by the date of diagnosis to identify the multimorbidity progressions each individual contributed prevalence to. Once all progression sets for all individuals had been processed, a set of observed, unique disease sets (hyperedges) and multimorbidity progressions (hyperarcs) were identified and their weights were computed. This was carried out for both the Charlson and Elixhauser populations.

The weights of each hyperedge corresponded to the proportion of individuals that progressed to have the diseases which the hyperedge connects, relative to all other similar diseases containing a subset of, or is a superset of, the disease set in question. The weight of a child hyperarc of this particular hyperedge is the proportion of individuals who were observed to have that exact disease progression out of all other possible disease progressions that the parent hyperedge can give rise to.

When constructing the PTM’s of the directed hypergraph, we utilised two transition rules to identify successor and predecessor conditions, the forward and backward transition rules. The forward transition rule follows the inherent direction of a hyperarc, and specifies a nonzero transition probability for any two disease nodes if there exists an observed multimorbidity progression which sees the source disease diagnosed *prior* to the target disease. The backward transition rule follows the opposite direction of a hyperarc, specifying a nonzero probability for any two diseases if there exists an observed multimorbidity progression which observes the source disease diagnosed *after* the target disease. Higher pairwise transition probabilities are observed for diseases which are sequential in nature and are highly prevalent among the population relative to other transitions of the same diseases.

Population-level estimates of the progression probability of reaching a target disease node, irrespective of the source disease, can be achieved by using PageRank centrality on these successor and predecessor PTMs. It is this centrality measure which is used to identify the temporal roles each disease plays in both populations, be it predecessory, successory or transitive. We then constructed directed hypergraphs and used PageRank on different strata for each sub-cohort, split by age, sex, age and sex, and socioeconomic status. To demonstrate the additional information captured by the directed hypergraph, a directed graph was constructed and PageRank was applied for comparison.

Sex was defined as the assigned gender at birth and socioeconomic status was defined by the 2011 Welsh index for multiple deprivation (WIMD2011) quintiles. WIMD2011 was based on linkage to the Lower-layer Super Output Area (LSOA) of an individuals place of residence at the time of entry into the study (on January 1st, 2000). Similarly, age was calculated as an individuals age upon entry into the study and was organised into 6 categories (20–39, 40–49, 50–59, 60–69, 70–79 and 80–99) — this was to ensure a sufficient proportion of the population sampled per age category. Age groups were mutually exclusive, such that no one participant could belong to more than one age group.

The directed hypergraph framework and PageRank analysis was implemented and carried out in Python (version 3.8.8). The code base used in this work is freely available on GitHub [45]. To deal with the computational resources required to construct the directed hypergraphs, we used the Python package Numba (version 0.56.3) [46]. This was particularly effective for data cleaning, edge weight computation and PageRank analysis. Numba is a just-in-time (JIT) compiler for the Python interpreter which converts basic syntax for array-based operations into machine code, resulting in comparable runtime performance with traditional compiler languages [46].

We had two machines available on SAIL, an AMD Epyc 7543 with 4 cores and 16Gb memory (The standard SAIL virtual machine), and one with more compute resources available, an Intel Xeon 6138 with 64 cores and 256Gb memory (made available via a JupyterHub server). While both are running on a server or workstation, the former is reasonably comparable to a standard desktop PC.

## 3 Results

There were 2,902,101 individuals in the WMC, reduced to 2,178,938 (75.1%) for the WMML cohort of individuals aged 20 or above [7]. Some 1,313,219 (45.3%) individuals had at least one condition in the Charlson comorbidity table, referred to as the Charlson population, while 1,709,340 (58.9%) individuals had at least one condition in the Elixhauser comorbidity table, referred to as the Elixhauser population. After excluding individuals with 3 or more conditions observed for the first time on the same date, we retained 1,298,601 (44.7%) and 1,639,695 (56.5%) individuals as our cohorts used in the analysis. See supplementary figure S4 for a flow chart on our analysis cohort specification for Charlson and Elixhauser populations. Supplementary tables S6 and S7 describe the population demographic statistics — split by disease — in both the Charlson and Elixhauser populations, respectively.

Table 1 reports progression duplicates, hyperedge and hyperarc information on the directed hypergraphs of the entire Charlson and Elixhauser populations, respectively. Due to their being many more conditions present in the Elixhauser comorbidity index, over half of the Elixhauser analysis cohort were multi-morbid. This is in contrast to the Charlson analysis cohort, where just under a third of the cohort were multi-morbid. The directed graphs are fully connected by virtue of directed edges only connecting two nodes, whereas the undirected and directed hypergraphs are not. In fact, in both populations their hypergraphs are empty relative to the maximum number of connections available. We also recorded the elapsed time taken to fully process all participants and construct the directed (and undirected) hypergraph models.

**Table 1:** Population and graph model statistics for Charlson and Elixhauser populations. For sample size statistics, percentages refer to proportion of total sample size available. For graph model statistics, percentages refer to proportion of maximum number of edges. The standard SAIL virtual machine to process the Elixhauser population due to a lack of available memory.

|  | Charlson population | Elixhauser population |
| --- | --- | --- |
| Number of disease nodes | 13 | 27 |
| Total population | 1,313,219 | 1,709,340 |
| Analysis cohort | 1,298,601 | 1,639,695 |
| 1-condition | 557,798 | 387,247 |
| 2-condition | 325,945 | 342,763 |
| $n$-condition ($n > 2$) | 414,858 | 909,685 |
| Unique hyperedges | 5,514 (67.3%) | 290,791 (0.2%) |
| Unique hyperarcs | 20,254 (38.0%) | 568,025 (0.03%) |
| Directed graph edges | 169 (100%) | 729 (100%) |
| Standard Runtime | 5.1 s ± 25.6 ms | - |
| JupyterHub Runtime | 4.25 s ± 33.4 ms | 2.05 minutes ± 954 ms |

Figure 2 shows the 28 top-weighted multimorbidity progressions which feature specific conditions for both the Charlson cohort (2a) and the Elixhauser cohort (2b). Subfigure 2a shows highly weighted progressions which feature myocardial infarction (MI) and congestive heart failure (CHF). Subfigure 2b shows highly weighted progressions where alcohol abuse and drug abuse appear as precursory conditions. To see the highest weighted progressions for both cohorts in their respective directed hypergraphs, see supplementary figure S5.

**Figure 2:**
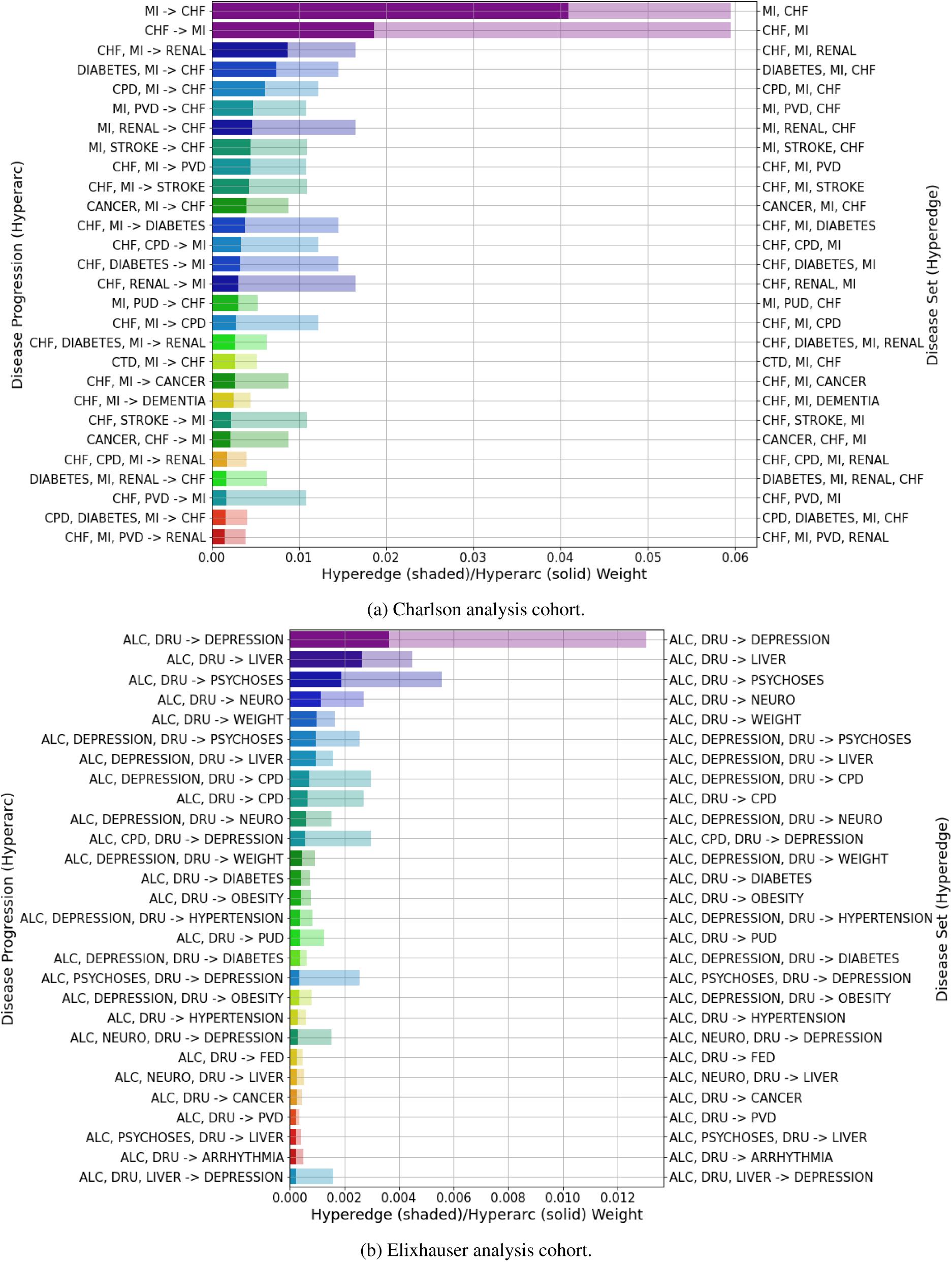
Top 28 hyperarc (solid) and hyperedge (shaded) weights which feature specific diseases for both populations. In (a) MI and CHF feature in the multimorbidity disease progression in the Charlson analysis cohort. In (b) alcohol abuse and drug abuse feature as precursory comorbidities in each multimorbidity disease progression. Here, ‘ALC’ and ‘DRU’ were used as shorthand for ‘ALCOHOL’ and ‘DRUG’.

Figure 2 presents a pattern in both cohorts, identifying conditions which act as precursors to other subsequent diagnoses, called successor conditions. For example, in subfigure 2a, we see MI act precursory to CHF, since *MI* → *CHF* is superior compared to its sibling *CHF* → *MI*. Moreover, most top-weighted progressions have CHF, renal disease and peripheral vascular disease (PVD) succeeding other conditions like diabetes and MI [47]. Similarly, in subfigure 2b, disorders like liver disease, psychosis and other neurological disorders (OND) appear as successors in the highly weighted hyperarcs. Moreover, depression appears as a major comorbidity coincident with alcohol and drug abuse.

These pairwise, temporal relationships can be quantified more rigorously by constructing a discrete-time Markov chain transition matrix for both successor and predecessor transition rules. Figure 3 shows the PTMs computed from both analysis cohorts, using forward as well as backward transition rules. The (*i, j*)^th^ probability values in each matrix, represented as heatmaps, correspond to the probability of condition *j* being a successor/predecessor of condition *i*.

**Figure 3:**
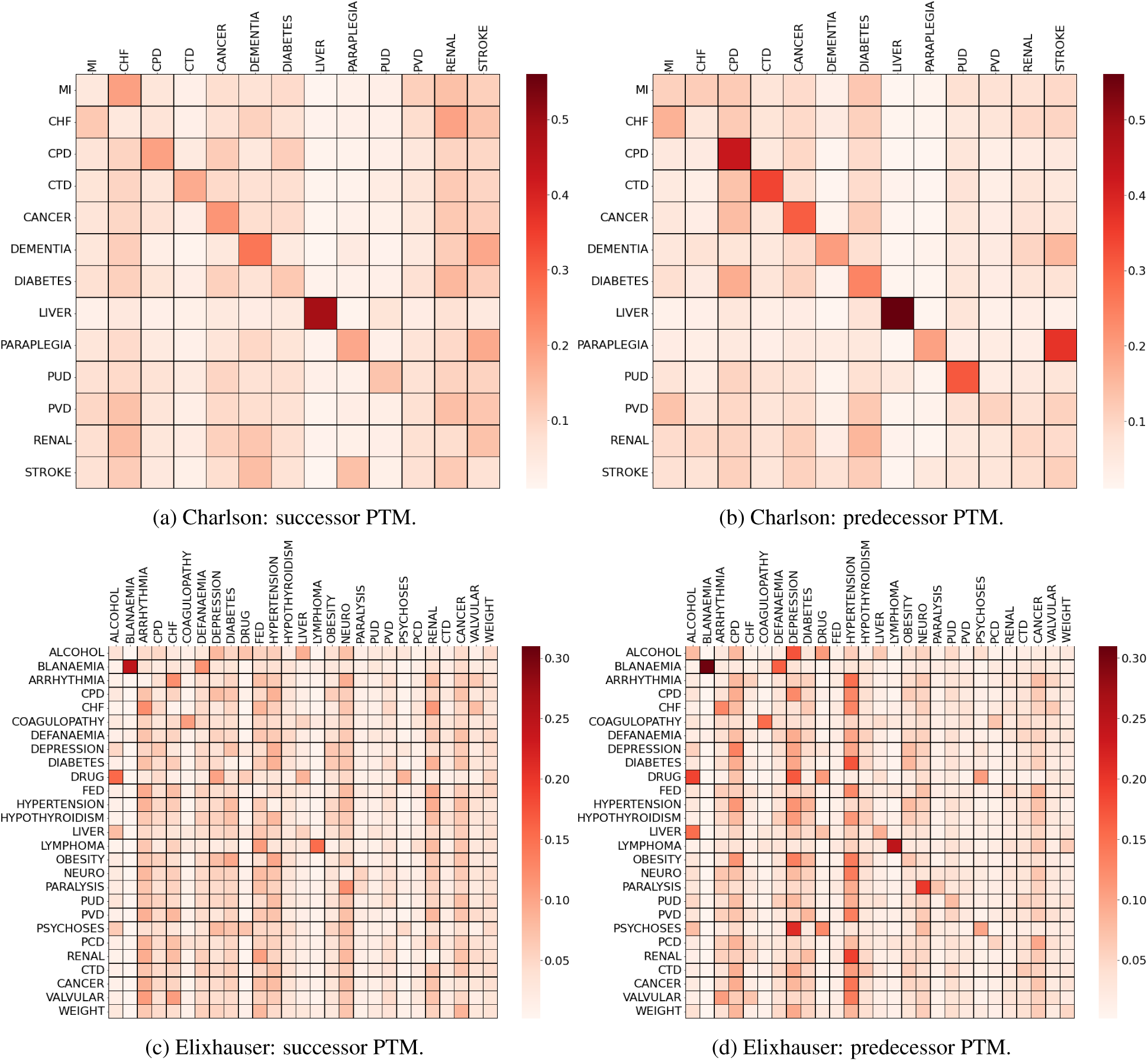
Probability transition matrices for the Charlson (a–b) and Elixhauser (c–d) analysis cohorts.

Figure 3 helps identify influential conditions from a temporal perspective for both successor and predecessor conditions. This can be done qualitatively by inspecting the values of each column of the PTMs. For example, in the successor PTMs in figures 3a and 3c we observe renal disease and arrhythmia to be influential successor conditions, while in the predecessor PTMs in figures 3b and 3d chronic pulmonary disorder (CPD) and hypertension have the strongest predecessor transition probabilities, respectively. These high values are contrasted by liver disease and blood loss anaemia in figure 3, which are not prominent predecessor or successor conditions with the remaining conditions in their respective comorbidity disease tables. This is because their prevalence is primarily made up from single-disease participants, hence inflating their diagonal transition probability entry, which necessarily has a downstream effect when using centrality metrics.

To visualise the combined role each disease plays in the population, we can plot the PageRank value of both transition matrices in two separate axis. Figure 4 present the successor and predecessor PageRank probabilities for both the directed hypergraph and directed graph case in each analysis cohort, respectively. These plots are helpful for identifying conditions which are significant successors, predecessors, both or none at all. To better understand the comparison with the directed hypergraph and simple directed graph case, supplementary figure S7 plot the signed difference computed between the predecessor and successor PageRank centrality values for both analysis cohorts.

**Figure 4:**
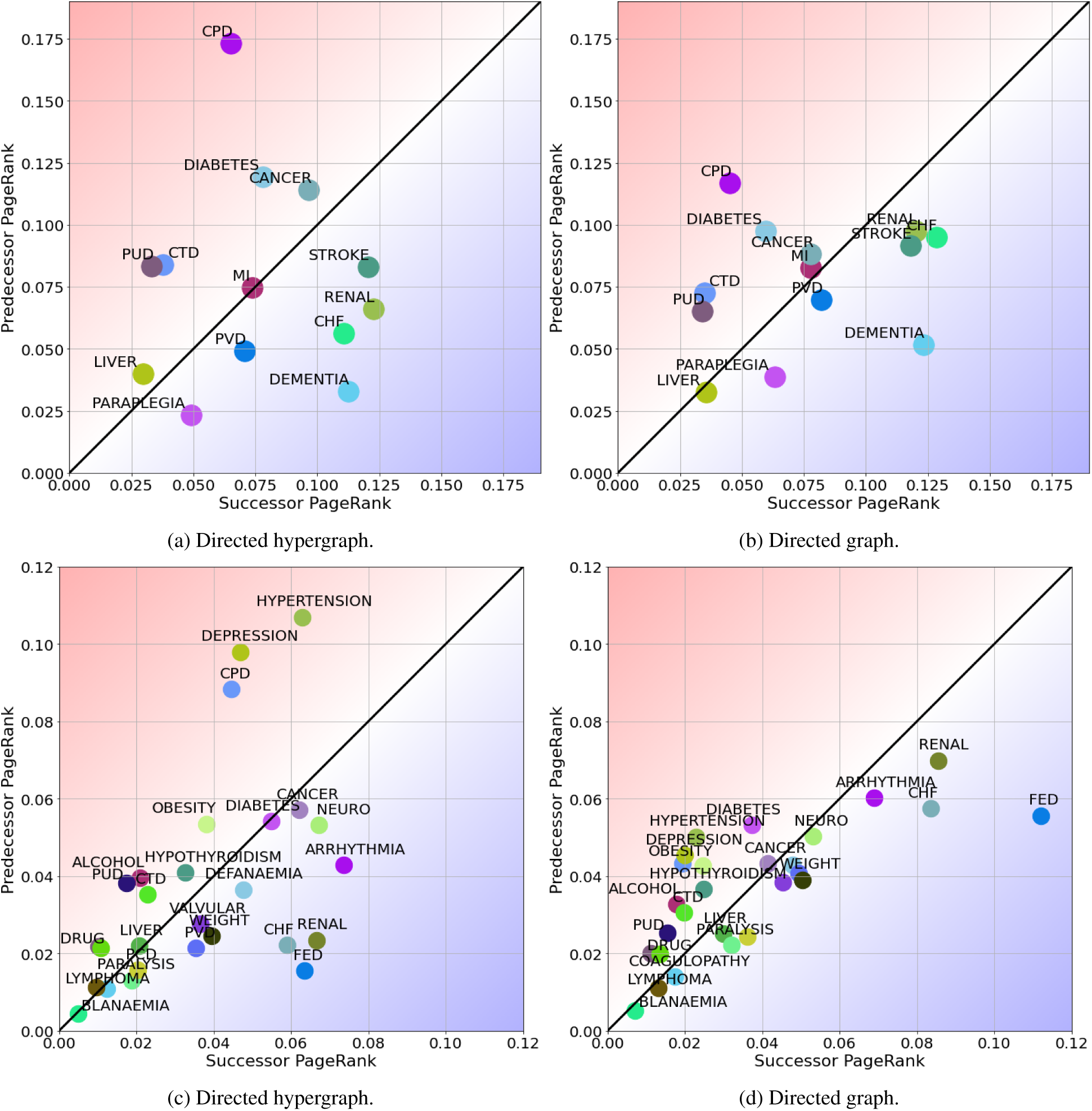
Visualisation of successor and predecessor PageRank values for conditions in Charlson (a–b) and Elixhauser (c–d) analysis cohorts using the directed hypergraph (a,c) and directed graph (b,d). In (c) titles for coagulopathy and psychoses are omitted as they overlapped with lymphoma and drug abuse, respectively. In (d) valvular disease, peripheral vascular disease and anaemia deficiency were all omitted as they overlapped with liver disease. Moreover, psychoses, CPD and PCD were omitted as they each overlapped with drug abuse, obesity and liver disease, respectively.

Figure 5 show the change in PageRank centrality ranking for successor and predecessor detection after stratifying each cohort by age group. Figures 6 show results from computing PageRank centrality for the Charlson and Elixhauser analysis cohorts stratified by sex. Line widths are shown where conditions have changed ranking across sexes and scatter plot diameters measure the population of each disease within the sex-stratified populations. PageRank centrality was also computed for combined age-sex demographic stratification’s. These results can be found in supplementary figures S8, S9. We also investigated stratification across all 5 deprivation indices but this data is not shown due to a lack of significant results.

## 4 Discussion

In this study, we introduced directed hypergraphs as a way to model multimorbidity development, a novel framework utilising multimorbidity prevalence to understand multimorbidity disease progression. To highlight its utility in the study of multimorbidity, we identified directional and contributory relationships among conditions in each comorbidity index. We constructed discrete-time Markov chain models to represent pairwise directional transitions between conditions and computed PageRank to assess the contributory relationships conditions had with one another. We believe this to be the first study conducted which looks at modelling disease transitions using directed hypergraphs and as such this study acts as an introduction to the directed hypergraph framework, allowing a diverse range of downstream tasks to be performed such as the inclusion of temporality and mortality.

### 4.1 Population-level results

Most of the top ranking hyperarcs of the Charlson analysis cohort were self-transition ones, as 43% of individuals had only one condition (see supplementary figure S5a). In contrast, the distribution of hyperarc weights for the Elixhauser analysis cohort showed the top prevalence-weighted hyperarcs represented comorbidity and multimorbidity rather than solitary conditions. This was due to the higher percentage of non-solitary disease progressions observed in the Elixhauser analysis cohort (76.4%). Furthermore, the applied weighting scheme penalised progressions with higher condition counts because an additional condition within a progression allows many more terms in the denominator of equation (1) to be added to the overall penalisation. This is also a realistic quantification of prevalence, as not everybody who had the progression *diabetes* → *renal disease* went on to develop acute MI, *diabetes, renal disease* → *MI*. Therefore, progressions with fewer condition counts should necessarily have a higher prevalence.

Figure 3 contain barplots that show how children hyperarc weights interact with their parent hyperedge weight, and thus, visualises the proportional difference between the clustering of diseases and the likelihood of progression between said diseases. For example, given the disease set {*CPD, cancer*}, the prevalence-weight of the multimorbidity progression *CPD* → *cancer* was larger than the alternate progression *cancer* → *CPD*. This prevalence superiority in the population is perhaps unsurprising given the relationship between CPD and lung cancer [48], as well as the fact that lung cancer is one of the four most dominant cancers in terms of incidence in Wales, alongside prostate, breast and bowel cancers [49].

The probabilities along the diagonal entries of the PTMs for the Charlson cohort were more pronounced compared to those in the Elixhauser cohort. In general for the successor PTM, the individual entries are the probability of the disease being the final or only one in a progression series, for example dementia and liver disease in figure 3a had high probabilities in the successor PTM. In the predecessor transition matrix, this is simply the probability that this condition was the only one diagnosed across the period of analysis, examples having high probabilities being CPD, cancer and liver disease.This also reflects the hyperarc-hyperedge weight plots we see in supplementary figure S5a, where these diseases in particular had large self-transition hyperarc weights.

These large self-transition probabilities may relate to fatal conditions, where patients are diagnosed and die shortly afterwards without time to acquire additional diseases — liver disease has seen a 500% rise in all-cause mortality since 1970, for example [50]. This observation more generally leads to an important conclusion that disease sets must be appropriately chosen for the research question under consideration. For example, in subfigures 3c and 3d we observed significant relationships drawn between liver disease and alcohol abuse, drug abuse and fluid and electrolyte imbalance (FED). Because the latter three conditions are not present in the Charlson comorbidity index, we could have been incorrectly led to believe in the solitary nature of liver disease when grouped together with mostly cardiac conditions in the Charlson PTM’s in subfigure 3a and 3b. Moreover, 76.4% of the Elixhauser analysis cohort were at least comorbid compared to 57% in the Charlson analysis cohort, so there were fewer individuals with self-disease transitions in the Elixhauser analysis cohort, as seen in table 1.

This highlights a potentially unfortunate effect of centrality measures which look at population estimates on the graph. Given the Charlson comorbidity table primarily contains cardiac-related conditions, the temporal roles of other non-cardiac conditions such as liver disease and peptic ulcer disease (PUD) are suppressed in the PageRank centrality results. This can therefore neglect conditions which could have serious risk factors and comorbidities with other diseases not present in comorbidity table. Conducting the same analysis on subsets of conditions may help mitigate this issue, or including more conditions which have known comorbidities with the other disease nodes in the graph. The latter can be observed when analysing the Elixhauser analysis cohort, where comorbidities of liver disease are present in the condition list.

The PageRank visualisations are helpful in identifying conditions which are primarily successors, predecessors, both or none at all. In the Charlson analysis cohort, diseases like CPD, PUD, connective tissue diseases and diabetes were more predecessor than successor in nature. This is in contrast to stroke, CHF, renal disease and dementia which were major successors. In the Elixhauser analysis cohort, we observe diseases like CPD, depression and hypertension as major predecessors and arrhythmia, CHF, renal disease and FED as major successors. It is tempting to conclude that diseases such as stroke and dementia are successors because they often result in mortality. This is not necessarily the case, as in our analysis a disease with a high probability in the successor PTM simply means it is likely to occur following other diseases.

There are certain diseases which behaved equally as a successor and a predecessor condition, defined by having similar successor and predecessor PageRank values. We denote these diseases as a bridge condition, acting as transitively to preceding and superseding diseases. For example, MI had almost identical successor and predecessor PageRank values in the Charlson analysis cohort. This suggests that some precursor conditions increase the risk of MI such as diabetes [51], while after the cardiac event, MI influenced or triggered the onset of other conditions in an individual’s lifetime such as CHF [47]. The presence of these bridging conditions could also be related to the consideration taken by the clinical team during treatment, whereby conditions like MI may receive more intense observation, and thus more chance of further successor conditions being detected, than other predecessor conditions. However, the absence of difference in successor-predecessor PageRank values could be due to a lack of prevalence and connectivity of the condition within the population. For example, in the Elixhauser analysis cohort, blood loss anaemia has very similar successor-predecessor PageRank values, but only 0.4% of individuals presented with this condition in the cohort. This is contrasted with 13.4% of individuals who presented with MI during their multimorbidity development in the Charlson analysis cohort.

#### 4.1.1 Directed hypergraph and graph

In this work we compared the PageRank centrality on the directed hypergraph with the directed simple graph approach. In the directed simple graph, there are many more highly weighted hyperarcs which have many high-scoring successor conditions as members of the tail. See supplementary figures S6a and S6b for hyperarc-hyperedge weight plots using the directed simple graph. This is because their prevalence is penalised by the prevalence of the two conditions as solitary conditions, which is typically unlikely because they are more likely to develop from other conditions, such as kidney disease after diabetes. Therefore, when constructing the probability transition matrices, the probability between one source successor condition to a target successor condition is necessarily higher than the same probability using the directed hypergraph.

This is because for the directed hypergraph, successor conditions are observed in the tail of a hyperarc for parent hyperedges which connect more than 2 conditions, and therefore they have lower weight than smaller disease sets according to equation (1). Therefore, the probability of the successor being a predecessor condition is smaller than for the directed graph, because we represent these successor conditions using higher order hyperarcs. Where successor conditions appear in hyperedges that connect two conditions, they are more likely to appear as the head in the hyperarc, and therefore as a successor, for these highly weighted hyperarcs. This is why we see so-called successor conditions rank higher than we might imagine in predecessor detection for the directed simple graph, and we see the opposite for the directed hypergraph. This is the benefit of being able to model multimorbidity using higher order representations through the directed hypergraph.

The impact this has can be seen in figure 4, where many of the conditions in the directed simple graph are clustered around the boundary separating successor from predecessor. In the directed hypergraph, conditions are dispersed across the space of predecessor and successor regions. This can also be observed from the signed difference plots in supplementary figure S7, where the distance between successor and predecessor PageRank values are greater in almost all conditions using the directed hypergraph. Moreover, the directed simple graph identifies diabetes as a significant predecessor, while the directed hypergraph produces similar successor and predecessor PageRank values, suggesting it may act as a transitive condition in multimorbidity development. In the Elixhauser comorbidity table, there are many conditions which could lead to and are common comorbidities of diabetes, such as hypertension, obesity and also alcohol abuse. Likewise, there are successor conditions which refer to diabetes as a major risk factor like CHF or renal disease. Therefore, while in the Charlson analysis cohort, Diabetes is seen as a major predecessor, this relationship changes in the directed hypergraph with the inclusion of risk factors for diabetes. This change was identified by the directed hypergraph by virtue of being able to model higher-order disease progressions rather than comorbid disease progressions like the directed simple graph.

### 4.2 Demographic-level results

In the Charlson population, dementia appears as a major successor but a poor predecessor as seen in subfigure 4a. This role is perhaps unsurprising because of relatively increased prevalence among older adults, and so the condition is more likely to appear toward the end of many progressions, relative to the other conditions in the Charlson comorbidity index. This may misrepresent successor conditions of younger individuals, where dementia is less common. Therefore, it’s important to understand the directional relationships among different demographic factors such as age, sex and deprivation status.

We found significant variation in major successor and predecessor conditions for different age groups (figure 5). In the Charlson analysis cohort, diabetes and cancer appeared as the major successors in the youngest age groups, but declined rapidly for older individuals. This is because as individuals age and become multi-morbid, more serious conditions such as CHF and renal disease succeed these precursor conditions; diabetes for example is a known risk factor for several conditions, including those in both Charlson and Elixhauser comorbidity indexes [52]. CPD, cancer and diabetes remained major predecessors throughout while in individuals younger than 60, CTD ranked no lower than fifth, which can be understood by its prevalence among younger adults [53]. Stroke remained an important successor condition across age groups, reaching its peak for individuals between 50 and 59 and decreasing in rank to fourth for individuals aged 80 to 99. Given that stroke also increased in predecessor rank for older age groups from age 60 onward, this suggests stroke instances are common successors between ages 50-70, with stroke seen as a major predecessor in stroke survivors from age 70 onward [54]. A similar conclusion can be made for CHF, and it is also unsurprising that dementia was the major successor in the oldest age group [55]. These analyses highlight the importance of patient-centred care and treatment planning, since previous successor conditions can become predecessor conditions.

**Figure 5:**
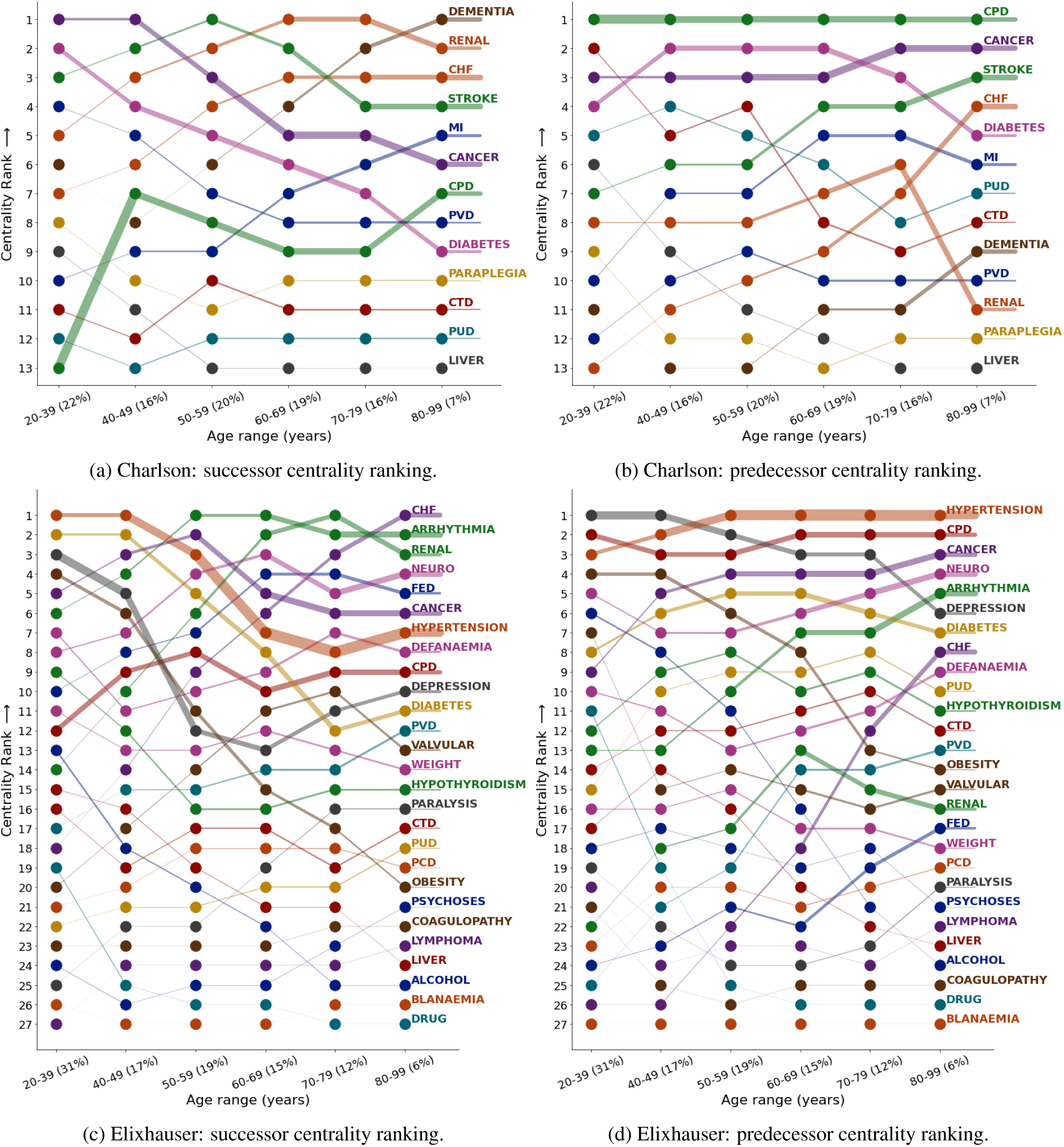
PageRank centrality ranking of diseases in the Charlson (a–b) and Elixhauser (c–d) cohorts stratified by age group. Age group proportion of population size are defined on the *x*-axis beside the age group title. The line width for each line segment connecting any two scatter points for a disease is proportional to the population size of that disease for the age group of the left-most scatter point. For example, in figure 5a, the disease with the largest population is CPD across all age groups. Disease labels on the right-hand *y*-axis represent the line next to it, rather than the centrality ranking.

In the Elixhauser cohort, we observed similar dynamics as in the Charlson cohort; diseases like diabetes, hypertension, obesity and depression appeared initially as major successors for younger age groups, before rapidly lowering in rank, remaining as major predecessor conditions throughout all age groups. These are common comorbidities with one another which were prevalent among younger populations (table S7) and are major risk factors for cardio-vascular conditions such as CHF, arrhythmia and renal disease. In contrast, and a striking similarity between both cohorts, was the appearance of CPD as a major predecessor consistently across all age groups while also remaining as a mid-to-low successor condition. Tables S6 and S7 show CPD was among the most prevalent conditions in the youngest age group and closely linked with other comorbidities like diabetes and hypertension [56]. This suggests that from a population perspective, CPD is not only well connected to many other conditions, but also plays a role in starting an individual’s multimorbidity development, given that other major predecessors like diabetes, hypertension and cancer are seen as successors in earlier age groups in figure 5.

It’s important to note that, particularly in the Elixhauser analysis cohort, there were many conditions which had low prevalence or were not as well connected and therefore did not appear to have any major successor or predecessor roles in the population. These conditions can be identified by the thin line-widths seen in figure 5, such as paraplegia or liver disease in the Charlson analysis cohort, and blood loss anaemia and drug abuse in the Elixhauser analysis cohort. PageRank centrality provides an estimate of the population-level, steady-state probabilities according to the graph weighting scheme and high connectivity. This leads us to note that these results are by no means related to any one individual’s multimorbidity development and will encourage common comorbidities to be ranked higher than uncommon ones. For example, liver disease plays a stronger role in the Elixhauser analysis cohort than the Charlson one due to the inclusion of common risk factors and comorbidities of liver disease, therefore increasing its connectivity.

The predecessor and successor diseases across different sexes are seen in figure 6. In the Charlson analysis cohort, dementia appeared as the major successor in females with renal disease ranked third, while in males renal disease was the major successor with dementia ranked fifth. This reflects the higher prevalence of dementia in females than males [57], while chronic kidney disease is more common in females but the treatment of end-stage kidney disease, dialysis and kidney replacement therapy is more common in males [58]. These differences were also observed in the successor condition rankings in the age-sex stratification presented in supplementary figures S8 and S9. CPD, cancer and diabetes remained strong predecessors for both sexes, while CTD was a stronger predecessor in females and MI in males, again aligning with the literature on these prevalence rates by sex [53, 51].

In the Elixhauser analysis cohort, there were consistent successor and predecessor conditions observed in both sexes and across most older age groups. Arrhythmia, neurodegenerative disorders, renal disease and FED were major successors while CPD, depression and hypertension remained as consistent predecessors. Note again, the emerging pattern of successors becoming predecessors in the PageRank centrality results of age-sex stratification in supplementary figures S8 and S9 as described above. Cancer for example ranked much higher as a successor condition in females than in males [49] (figure 6). In the predecessor PageRank centrality ranking, hypothyroidism and deficiency anaemia were stronger predecessors in females than in males in both sex and age-sex stratification’s. Compared with males, iron deficiency is relatively common in females due to the interaction between thyroid and menstrual hormones in younger females [59], and particularly iron deficiency anaemia among pregnant mothers [60] — two conditions observed as strong successors in the youngest age group of females (supplementary figure S9c).

**Figure 6:**
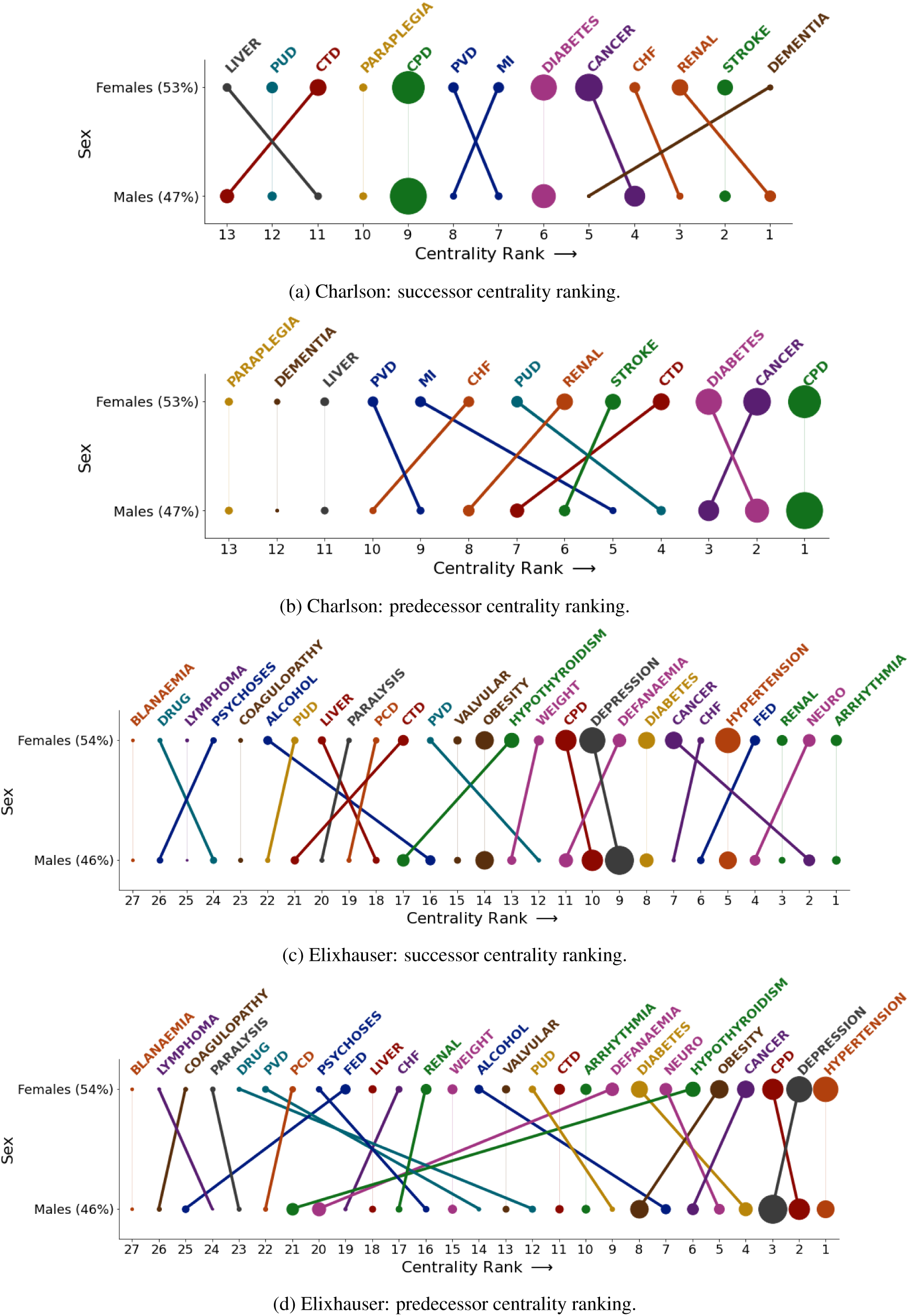
PageRank centrality ranking of diseases in the Charlson (a–b) and Elixhauser (c–d) analysis cohorts stratified by sex. Disease labels on the top *x*-axis represent the line next to it, rather than the centrality ranking.

We also investigated how PageRank centrality rankings changed according to the socioeconomic status using WIMD2011 quintiles. However, we saw little to no change in the roles diseases had as predecessors or successors. This is possibly due to how the WIMD2011 quintiles are calculated, which incorporates a health domain including all-cause mortality rate, cancer incidence and long-term illness [61]. Moreover, we observed the most dynamic change in successor and predecessor roles in the age stratified PageRank results, and since there was an even split across age (and sex) within each deprivation quintile, this may also explain why there is an approximately constant ranking across quintiles.

### 4.3 Future work

There are numerous ways the directed hypergraph approach could be extended and investigated further with respect to multimorbidity, with this study laying the foundations. At the heart of graph models are the weights which quantify the different relationships that are formed. These weights dictate the outcome and interpretation of the analyses. In our hyperedge weight formulation, the arbitrary coefficients in equation (1) could be guided with suitable domain knowledge on the chronic conditions being studied so that interaction effects of particular disease combinations can be integrated into the network directly through the edge weights. Moreover, while prevalence-weighted multimorbidity is a valid analysis pathway, other healthcare-oriented outcomes and mathematical formulations are equally feasible. For example, one could consider the healthcare resource ‘cost’ associated with a particular multimorbidity progression and perform a similar analysis, looking at how multimorbidity development trends differ between inpatient and outpatient care and what impact that has on available resources in the healthcare service.

From a technical perspective, we could look at other hypergraph-based approaches to model multimorbidity development. For example, instead of the directed hypergraph, we could consider an ordered, undirected hypergraph [62] which may provide a stronger representation of individual-based progression since here the ordering of tail sets in hyperarcs are preserved, while in our directed hypergraph the ordering is redundant. Alternatively, we could introduce more complex weighting systems such as edge-dependent node weights [63] to more accurately represent multimorbidity disease progression, driven by suitable domain knowledge.

Rafferty, et al. [30] discussed the concept of the dual undirected hypergraph which swapped the roles of the nodes and hyperedges in the graph, allowing them to assess eigenvector centrality on multimorbidity disease sets. A similar idea exists within the directed hypergraph space, which could allow analytical techniques like PageRank centrality to be performed on the multimorbidity disease progressions, rather than on the individual conditions. This could give us a way of quantifying how important specific multimorbidity trajectories are within the population.

At present, our directed hypergraph only models observable progressions of disease and there is no inclusion of mortality which does not reflect clinical progression accurately. It would be an interesting extension to include the explicit time between observed diagnoses within multimorbidity development, as well as the inclusion of finality into the directed hypergraph.

Because centrality is a population-level measure, it might also be appropriate to consider individual-based analyses, such as reconstructing the directed hypergraph to be used within a multi-state model. This could integrate demographic and lifestyle factors into the modelling process, rather than rely on stratification, which would allow these additional factors to influence the weighting scheme directly.

A disadvantage of hypergraphs in comparison to their directed graph equivalent is the difficulty in visualising the hypergraph network and their multi-way relationships. A publicly available applet to help visualise the undirected and directed hypergraph models using synthetic data has been built [64], allowing individuals to dive into the methodology from a visual perspective, helping engage with those less experienced with graph methods in the clinical space.

### 4.4 Limitations

There were a number of limitations in this work. No long-term, longitudinal study of multimorbidity is without right- and left-censoring. Most longitudinal multimorbidity cohorts endeavour to recruit individuals from birth and track their multimorbidity development until death. However, in practice datasets recruit some individuals only from a later stage in their life and will also undoubtedly track participants who are still alive by the end of the follow-up period and at risk of developing further conditions in their multimorbidity development. In the former case, historical records must be collected and linked while in the latter, an individuals multimorbidity development must be truncated for analysis. These cases can introduce a left- and right-censoring issue, depending on the quality of historic health records and the impact of incomplete multimorbidity trajectories. However, the WMC boasts a 20 year follow-up coverage to mitigate these issues.

Our analysis excluded participants who had three or more conditions observed during the same interaction with the healthcare system. While this only represented a very small proportion of individuals in both Charlson and Elixhauser populations, these individuals represent situations where chronic health conditions are consequential findings after a more serious event has been observed — such as the discovery of diabetes and renal problems after an acute MI. Removing these individuals with complex healthcare interactions may have introduced some selection bias into our analysis. Moreover, all multimorbidity progressions modelled from each individual were only *observed* from an EHR perspective, and can only approximate the real-life disease progression each participant went through, and it’s very likely some diagnoses were observed after the fact.

There is some disagreement over the exact definition of multimorbidity and many consider the term multimorbidity to encapsulate both *acute* and *chronic* conditions [1]. Our study, like most others, assumes this and considers multimorbidity development as an irreversible, one-way process. However, allowing acute illness to fall under the same branch of multimorbidity as chronic illness may be misguided. While an acute event can be justifiably argued to have long-lasting impacts to the body such as a MI or an acute stroke, these conditions can be considered *episodic* in nature. Moreover, these events can happen multiple times in ones life time, for example it is far more likely for another stroke to occur if an individual has already had one before.

During our analysis we identified disease nodes within the Charlson and Elixhauser comorbidity indices which are indeed acute, episodic events. For example, MI in Charlson, and alcohol abuse, drug abuse, obesity, weight loss and more in Elixhauser. Some of these conditions could even be considered entirely curable if caught and remedied. Modelling episodic events as well as chronic ones during an individuals disease trajectory could improve our understanding of the interaction effects between multiple, repeating acute events and singular, chronic events in ones lifetime. It could also reveal patterns from a healthcare resource perspective, as we would expect individuals with reoccurring acute episodes such as MI to be considered a priority for frequent check-ups. However, to implement this in practice would require time-intensive investigations into patient extracts to work out reliable episode number estimates, and would likely also add computational cost to constructing the directed hypergraph.

Constructing a hypergraph model is notably more computationally expensive than a simple graph. A fully connected, simple graph with 20 nodes has 190 connections, while there are over 1 million in a fully connected hypergraph. Matters are made significantly worse for a directed hypergraph, with over 10 million possible B-hyperarcs (and over 3.5 billion hyperarcs with arbitrary tail and head sets). While the hypergraph framework is ideal for representing multimorbidity, the number of possible connections permitted by an undirected hypergraph grows exponentially with the number of nodes, due to the many possible permutations of multi-disease sets. This is exacerbated even further by imposing directionality among the hyperedges. We note all of these issues are reflections of the complexity of the problem under consideration, and our current implementation takes about as much time to look outside and assess the weather to process the Charlson population, and the Elixhauser population as much time to boil a kettle — that is, to process over 1.6 million individuals with 27 disease nodes, extract their multimorbidity trajectories, compute hyperedge and hyperarc weights, build the directed hypergraph and carry out PageRank analysis. Ending on a note of caution — applying our current approach for a larger number of nodes may not be so forgiving in terms of processing time, but we are working on this.

## 5 Conclusions

In this work, we have established a new framework, the directed hypergraph, to model multimorbidity development and demonstrated its use case and utility with a large, longitudinal cohort using two standardised comorbidity index tables, the Charlson and Elixhauser. Patient disease progressions were modelled and quantified using relative prevalence and we have introduced a novel way to identify and visualise the temporal successor and predecessor roles conditions play among different demographics at a population level using PageRank centrality. We have also shown our analytical approach aligns with current literature on the contributory relationships between common comorbidities within different demographic strata and illustrated it’s power over simpler, directed graphical models. This is by virtue of it’s ability to model and quantify higher-order relationships over it’s simpler counterpart.

This preliminary study shows the use-case of directed hypergraphs in longitudinal multimorbidity analysis, and facilitates a number of new hypergraph-based approaches to emerge in multimorbidity investigation. We have released an open source implementation of the directed hypergraph to promote this further [45]. We encourage researchers in the multimorbidity community to extend and contribute to this work through the various pathways of analysis ranging from exploring different quantification’s of higher-order relationships, like healthcare resource utilisation, or restructuring the graph structure to be used within a multi-state model or multi-level Bayesian network to incorporate demographic information directly into the modelling procedure. Our team are currently investigating the inclusion of mortality and explicit temporality into the directed hypergraph. We hope in the future that the directed hypergraph can play an important role in multimorbidity investigation at both the population- and individual-level, helping guide the implementation of new healthcare strategies to improve quality of care and mitigate the risk of mortality of the most prevalent multimorbidity disease progressions.

## Data Availability

The data used in this study are available in the SAIL Databank at Swansea University, Swansea, UK. All proposals to use SAIL data are subject to review by an independent Information Governance Review Panel (IGRP). Before any data can be accessed, approval must be given by the IGRP. The IGRP carefully considers each project to ensure the proper and appropriate use of SAIL data. When approved, access is gained through a privacy-protecting trusted research environment (TRE) and remote access system referred to as the SAIL Gateway. SAIL has established an application process to be followed by anyone who would like to access data via SAIL https://www.saildatabank.com/application-process. This study has been approved by the IGRP as project 1392.

## 6 Acknowledgements

The core part of this work was completed as part of a Digital Analytics and Research Team (DART) PhD internship placement, within the NHS Transformation Directorate at NHS England. This study is based on work originally funded by the Medical Research Council (MRC) (Grant No.: MR/S027750/1) and supported by Health Data Research UK (Grant No.: HDR-9006), which receives its funding from the UK Medical Research Council, Engineering and Physical Sciences Research Council, Economic and Social Research Council, Department of Health and Social Care (England), Chief Scientist Office of the Scottish Government Health and Social Care Directorates, Health and Social Care Research and Development Division (Welsh Government), Public Health Agency (Northern Ireland), British Heart Foundation (BHF) and the Wellcome Trust; and Administrative Data Research UK, which is funded by the Economic and Social Research Council (Grant No.: ES/S007393/1). This study makes use of anonymised data held in the SAIL Databank, which is part of the national e-health records research infrastructure for Wales. We would like to acknowledge all the data providers who make anonymised data available for research.

## 7 CRediT authorship contribution statement

Jamie Burke: Methodology, Software, Formal analysis, Writing – original draft. Ashley Akbari: Writing – review & editing. Rowena Bailey: Writing – review & editing. Kevin Fasusi: Writing – review & editing. Ronan A. Lyons: Writing – review & editing. Jonathan Pearson: Writing – review & editing. James Rafferty: Supervision, Writing – review & editing. Daniel Schofield: Project administration, Supervision, Writing - review & editing

## 10 Appendices

### 10.1 Synthetic example

Table S1 shows the observed disease progressions for a synthetic dataset of 10 individuals with 3 disease nodes, *A, B* and *C*. Figure S1 plots the hyperedge and hyperarc weights for this synthetic dataset and figure S2 displays the weighted, directed hypergraph generated from this dataset. The hyperarc weights are represented visually through the thickness of the edges.

**Table S1:**
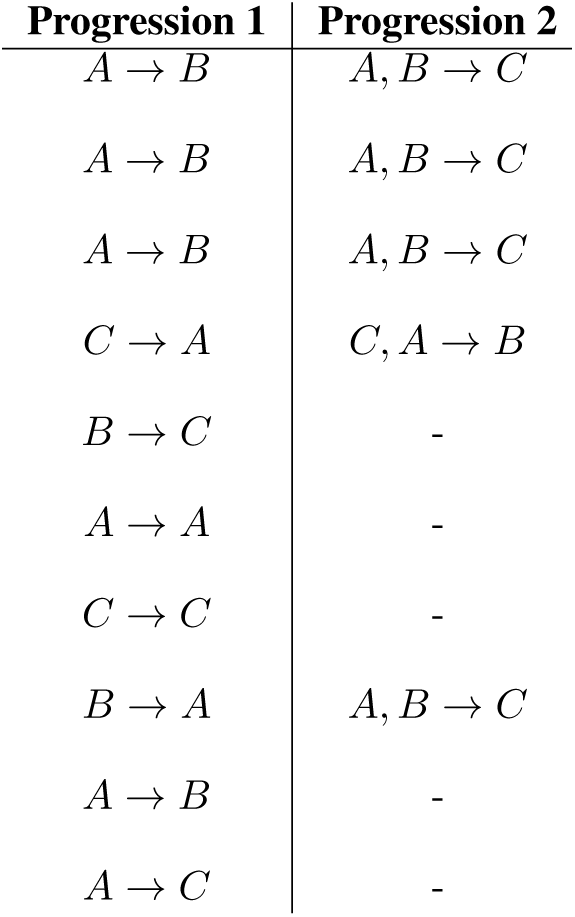
Progressions from synthetic data.

**Figure S1:**
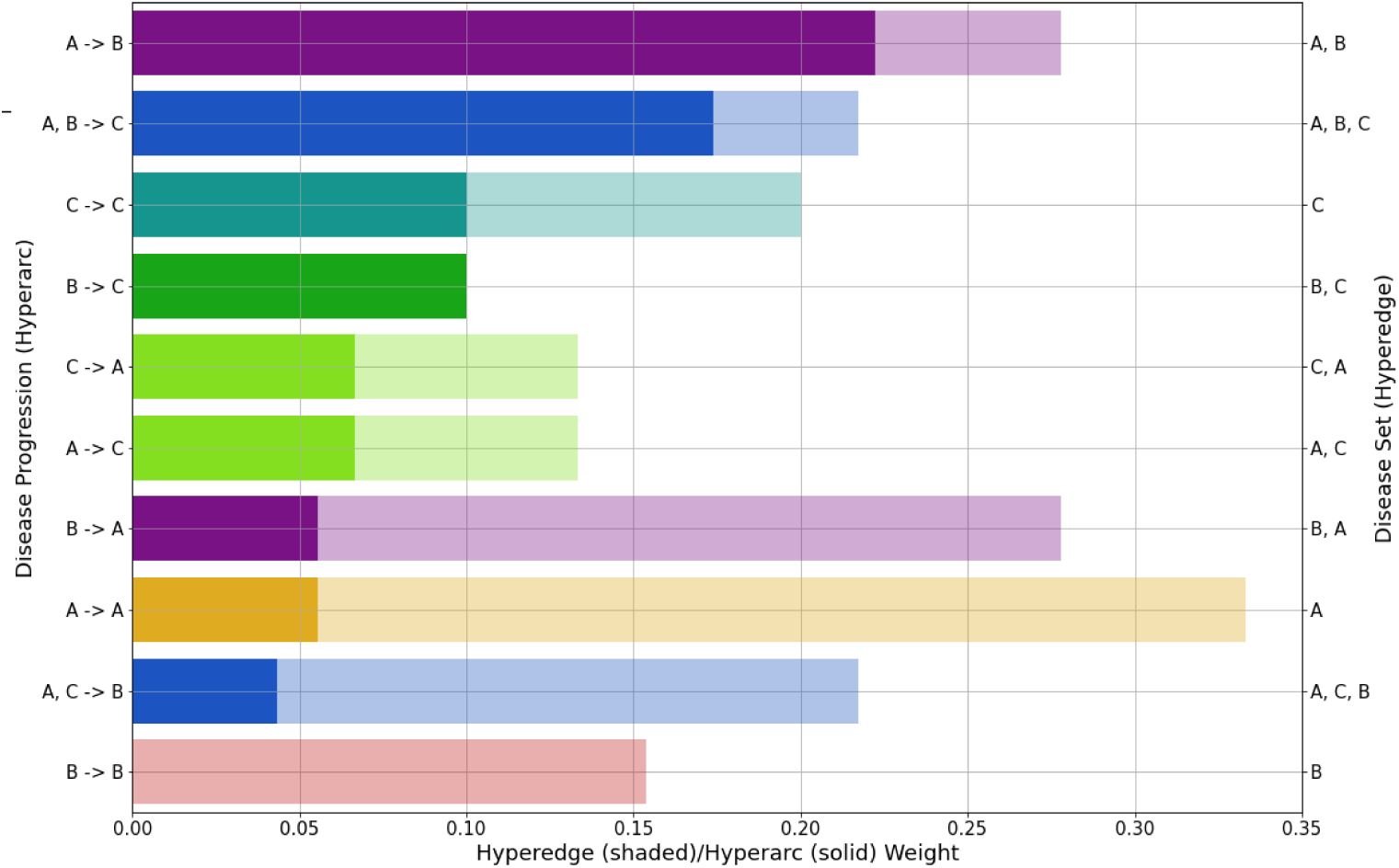
Hyperedge and hyperarc weights from synthetic data. Hyperarc weights (solid) are superimposed onto their corresponding parent hyperedge weight (shaded). Self-transition hyperarcs represent individuals observed with only that condition.

**Figure S2:**
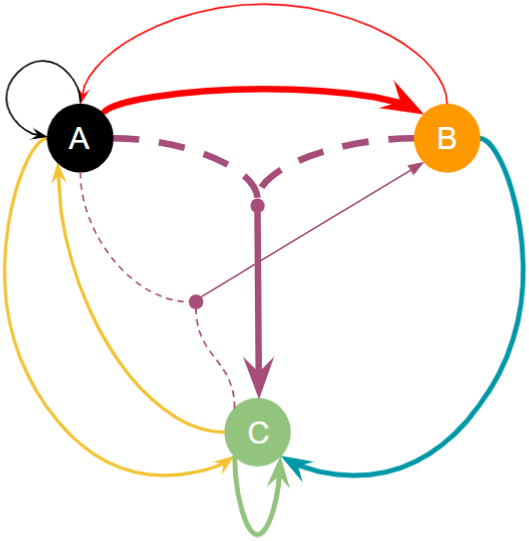
Weighted, directed hypergraph from the synthetic dataset described in table S1.

Figures S3(a-b) show the PTM’s for successor and predecessor transition rules. Computing the PageRank vector on both PTM’s and plotting each PageRank value on a different axis visualises the temporal role that a node plays in the directed hypergraph. In the synthetic dataset, the observed multimorbidity progression and subsequent PageRank values show the predecessor and successor nature of diseases *A* and *C*. Disease *B* has similar PageRank values for both transition rules, and is referred to as a *transitive* condition. This is a disease which is equally a predecessor as it is a successor, acting as an intermediary condition during an individual’s multimorbidity development.

**Figure S3:**
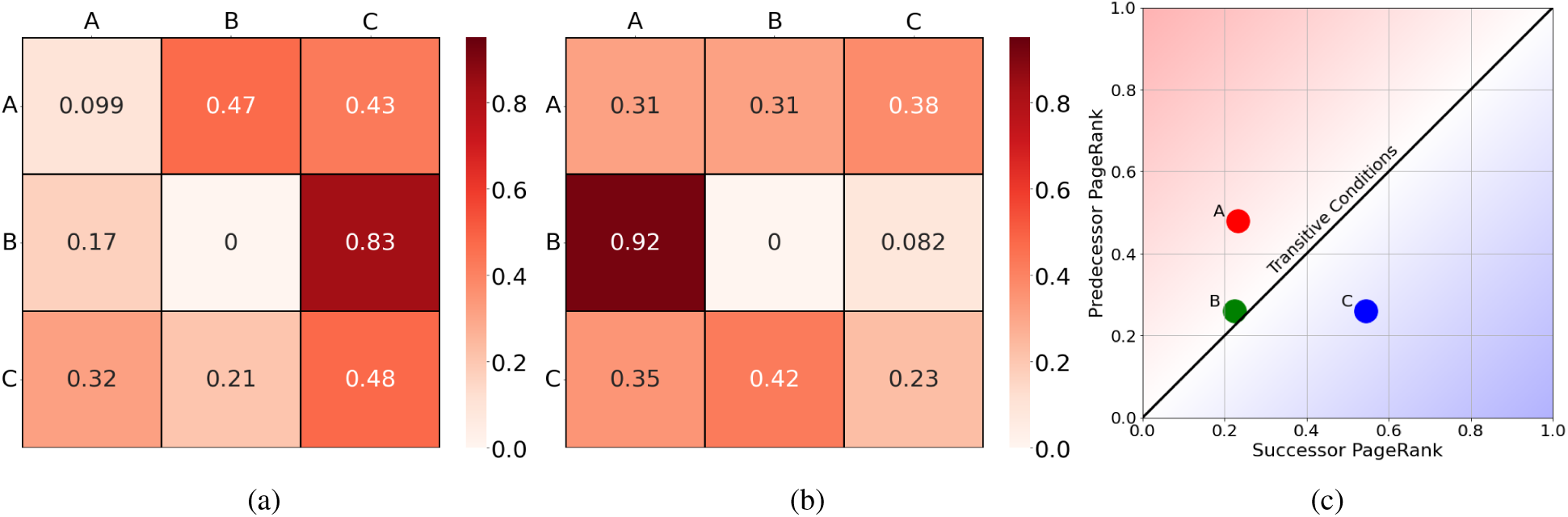
(a-b) Probability transition matrices for detecting successor and predecessor conditions on synthetic dataset. (c) Plotting successor and predecessor PageRank values on horizontal and vertical axes, respectively.

### 10.2 Comorbidity index disease tables

**Table S2:** List of comorbidities in the Charlson comorbidity index [41].

| Charlson Comorbidity Index (CCI) |  |
| --- | --- |
| Chronic Pulmonary Disease | Congestive Heart Failure |
| Connective Tissue Disease | Dementia |
| Diabetes (complication) | Diabetes (w/o complication) |
| Cancer, Lymphoma and Leukaemia | Liver Disease (mild) |
| Liver Disease (severe) | Metastatic Cancer |
| Myocardial Infarction | Paraplegia |
| Peptic Ulcer Disease | Peripheral Vascular Disease |
| Renal Failure | Stroke |

**Table S3:** CCI diseases and their corresponding title used for data visualisation. This also includes which conditions were combined into a single title.

| CCI Look-up Table |  |
| --- | --- |
| Chronic Pulmonary Disease | CPD |
| Congestive Heart Failure | CHF |
| Connective Tissue Disease | CTD |
| Dementia | DEMENTIA |
| Diabetes (w/ and w/o complication) | DIABETES |
| Liver Disease (mild and severe) | LIVER |
| Metastatic Cancer, Lymphoma and Leukaemia | CANCER |
| Myocardial Infarction | MI |
| Paraplegia | PARAPLEGIA |
| Peptic Ulcer Disease | PUD |
| Peripheral Vascular Disorders | PVD |
| Renal Failure | RENAL |
| Stroke | STROKE |

**Table S4:**
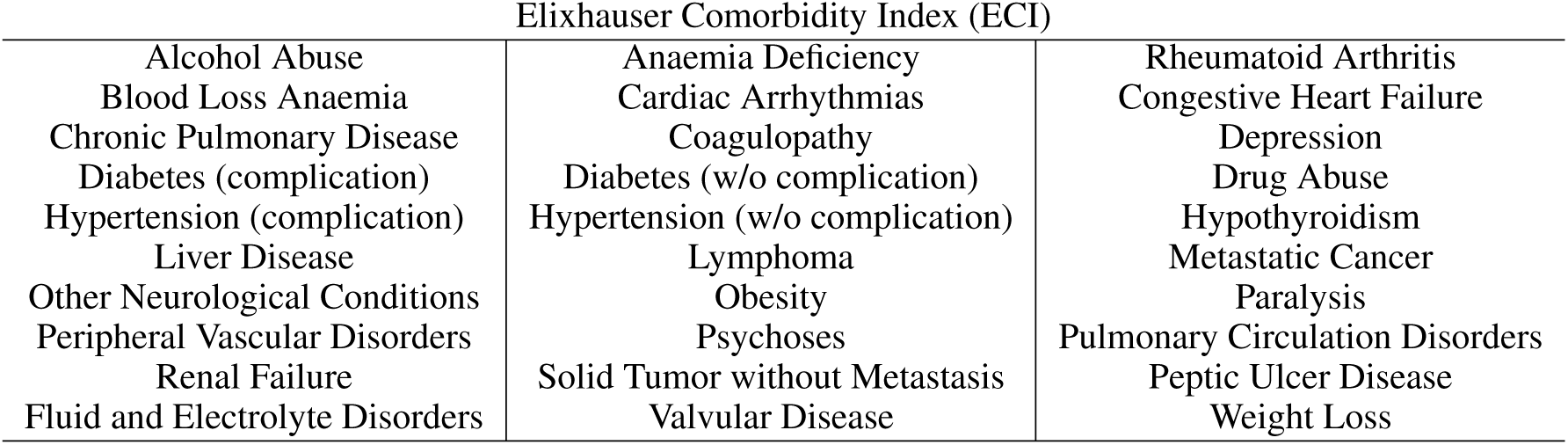
List of comorbidities in the Elixhauser comorbidity index [43].

**Table S5:** ECI diseases and their corresponding title used for data visualisation. This also includes which conditions were combined into a single title.

| ECI Look-up Table |  |
| --- | --- |
| Alcohol Abuse | ALCOHOL |
| Anaemia Deficiency | DEFANAEMIA |
| Rheumatoid Arthritis | CTD |
| Blood Loss Anaemia | BLANEAMIA |
| Cardiac Arrhythmia | ARRHYTMIA |
| Congestive Heart Failure | CHF |
| Chronic Pulmonary Disease | CPD |
| Coagulopathy | COAGULOPATHY |
| Depression | DEPRESSION |
| Diabetes (w/ and w/o complication) | DIABETES |
| Drug Abuse | DRUG |
| Hypertension (w/ and w/o complication) | HYPERTENSION |
| Hypothyroidism | HYPOTHYROIDISM |
| Liver Disease | LIVER |
| Lymphoma and metastatic cancer | LYMPHOMA |
| Solid Tumour w/o Metastasis | CANCER |
| Other Neurological Conditions | NEURO |
| Obesity | OBESITY |
| Paralysis | PARALYSIS |
| Peripheral Vascular Disorders | PVD |
| Psychoses | PSYCHOSES |
| Pulmonary Circulation Disorder | PCD |
| Peptic Ulcer Disease | PUD |
| Renal Failure | RENAL |
| Fluid and Electrolyte Disorders | FED |
| Valvular Disease | VALVULAR |
| Weight Loss | WEIGHT |

### 10.3 Population flowchart

**Figure S4:**
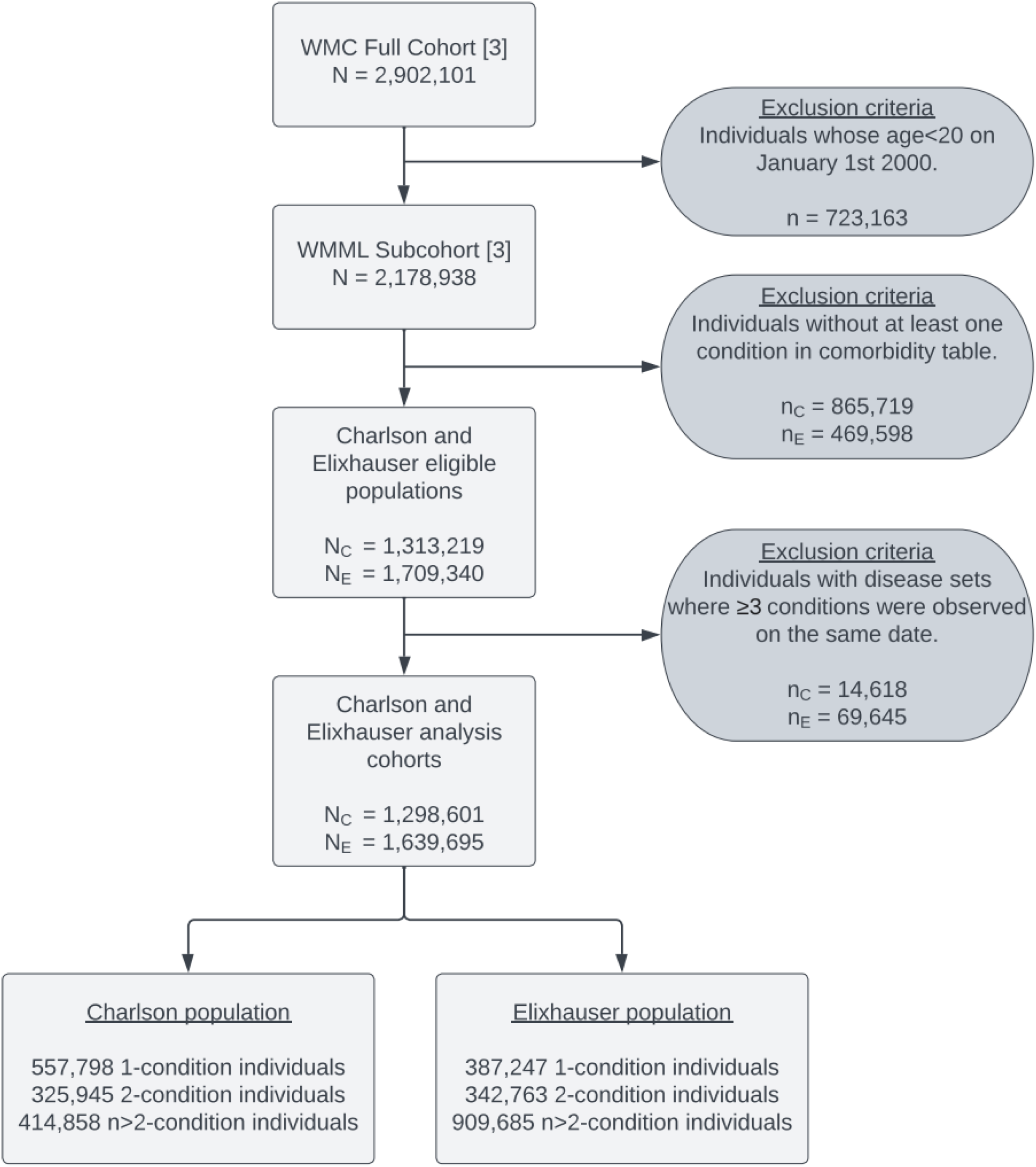
Flow chart visualising Charlson and Elixhauser cohort specifications. Upper-case *N* refers to sample size at each stage of specification and lower-case *n* refers to number of individuals excluded at each level. Subscripts *C* and *E* refer to Charlson and Elixhauser populations, respectively. WMC, Wales Multimorbidity e-Cohort; WMML, Welsh Multimorbidity Machine Learning [7].

### 10.4 Population and demographic statistics

**Table S6:** Population demographic statistics for Charlson population. Full disease names can be found via the look-up table in table S3.

| Disease | Proportion (%) |  |  |  |  |  |  |  |  |  |  |  |
| --- | --- | --- | --- | --- | --- | --- | --- | --- | --- | --- | --- | --- |
|  | Sex<br>(Males) | Age |  |  |  |  |  | WIMD |  |  |  |  |
|  |  | 20-39 | 40-49 | 50-59 | 60-69 | 70-79 | 80-99 | 1 | 2 | 3 | 4 | 5 |
| All Diseases | 52.6 | 21.9 | 16.0 | 20.2 | 18.5 | 15.9 | 7.5 | 21.1 | 20.6 | 20.6 | 18.8 | 18.9 |
| MI | 61.3 | 5.88 | 10.41 | 19.59 | 26.86 | 26.64 | 10.61 | 22.57 | 21.62 | 20.57 | 18.30 | 16.94 |
| CHF | 49.5 | 3.68 | 6.55 | 15.31 | 26.39 | 31.20 | 16.86 | 21.59 | 21.33 | 20.93 | 18.75 | 17.39 |
| CPD | 46.9 | 29.13 | 15.84 | 19.12 | 17.40 | 13.51 | 5.01 | 23.86 | 21.51 | 20.22 | 17.47 | 16.94 |
| CTD | 33.1 | 15.27 | 15.47 | 22.53 | 22.65 | 17.68 | 6.41 | 20.13 | 19.37 | 21.46 | 19.53 | 19.51 |
| CANCER | 48.7 | 9.90 | 13.84 | 24.09 | 25.02 | 19.80 | 7.34 | 18.74 | 19.66 | 20.69 | 19.72 | 21.19 |
| DEMENTIA | 36.8 | 1.14 | 2.35 | 9.00 | 26.62 | 39.72 | 21.18 | 20.13 | 20.59 | 20.74 | 19.06 | 19.48 |
| DIABETES | 51.4 | 18.80 | 17.42 | 23.33 | 21.53 | 14.42 | 4.49 | 23.10 | 21.31 | 20.61 | 17.53 | 17.45 |
| LIVER | 56.8 | 26.16 | 23.51 | 24.20 | 16.58 | 7.98 | 1.57 | 25.97 | 21.50 | 19.69 | 16.72 | 16.12 |
| PARAPLEGIA | 49.7 | 13.84 | 10.97 | 16.20 | 22.63 | 25.08 | 11.28 | 23.80 | 21.59 | 20.46 | 17.81 | 16.34 |
| PUD | 57.7 | 13.22 | 14.84 | 21.22 | 23.55 | 19.81 | 7.36 | 24.10 | 21.61 | 20.19 | 17.43 | 16.67 |
| PVD | 61.2 | 6.86 | 11.69 | 21.55 | 27.42 | 24.26 | 8.22 | 23.26 | 21.33 | 20.55 | 18.02 | 16.83 |
| RENAL | 46.2 | 6.11 | 9.07 | 20.05 | 29.24 | 27.11 | 8.43 | 21.37 | 21.01 | 21.28 | 19.15 | 17.19 |
| STROKE | 48.0 | 6.14 | 9.20 | 18.41 | 26.21 | 27.21 | 12.83 | 21.08 | 20.44 | 20.45 | 18.60 | 19.43 |

**Table S7:**
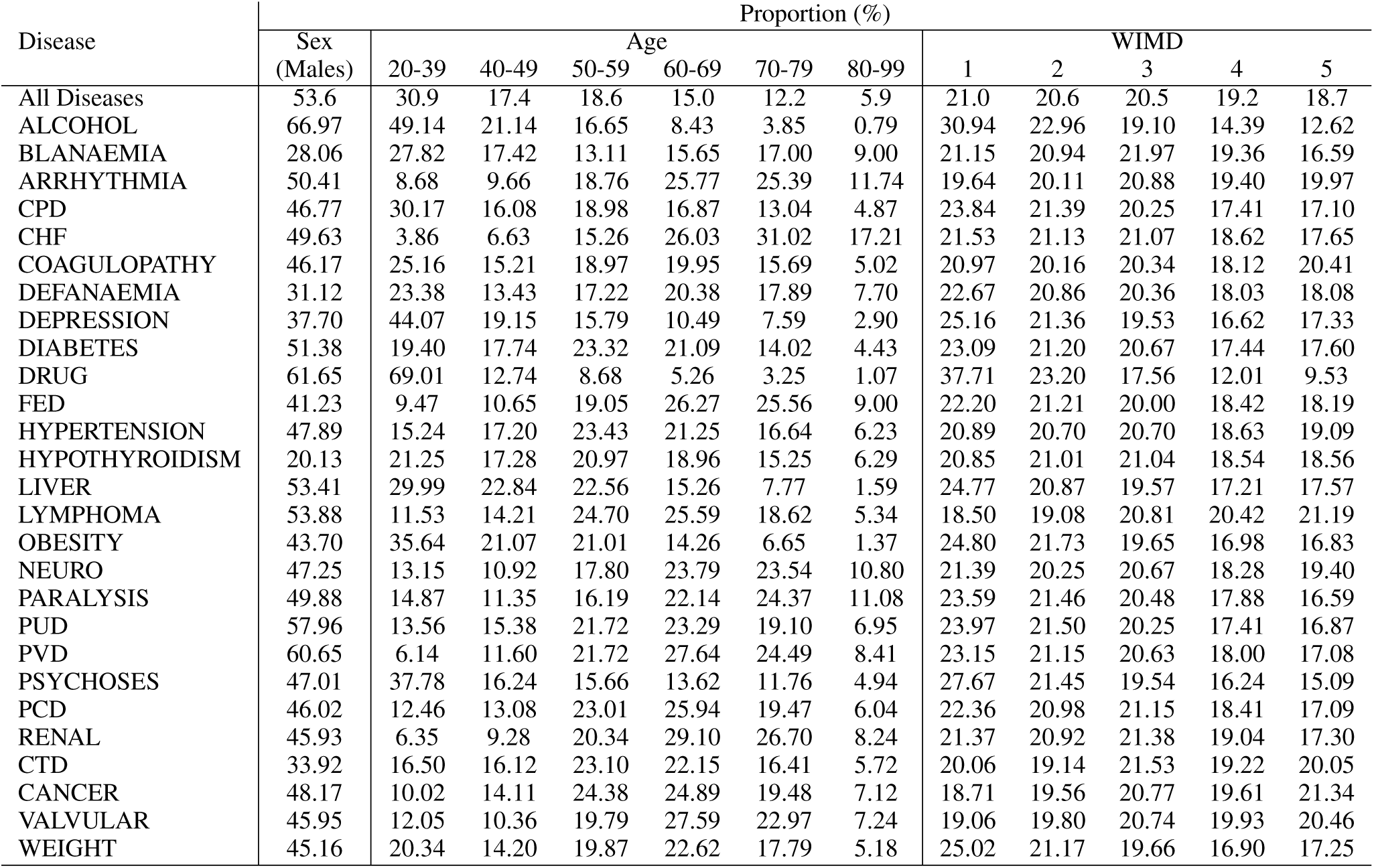
Population demographic statistics for the Elixhauser population. Full disease names can be found via the look-up table in table S5.

| Disease | Proportion (%) |  |  |  |  |  |  |  |  |  |  |  |
| --- | --- | --- | --- | --- | --- | --- | --- | --- | --- | --- | --- | --- |
|  | Sex<br>(Males) | Age |  |  |  |  |  | WIMD |  |  |  |  |
|  |  | 20-39 | 40-49 | 50-59 | 60-69 | 70-79 | 80-99 | 1 | 2 | 3 | 4 | 5 |
| All Diseases | 53.6 | 30.9 | 17.4 | 18.6 | 15.0 | 12.2 | 5.9 | 21.0 | 20.6 | 20.5 | 19.2 | 18.7 |
| ALCOHOL | 66.97 | 49.14 | 21.14 | 16.65 | 8.43 | 3.85 | 0.79 | 30.94 | 22.96 | 19.10 | 14.39 | 12.62 |
| BLANAEMIA | 28.06 | 27.82 | 17.42 | 13.11 | 15.65 | 17.00 | 9.00 | 21.15 | 20.94 | 21.97 | 19.36 | 16.59 |
| ARRHYTHMIA | 50.41 | 8.68 | 9.66 | 18.76 | 25.77 | 25.39 | 11.74 | 19.64 | 20.11 | 20.88 | 19.40 | 19.97 |
| CPD | 46.77 | 30.17 | 16.08 | 18.98 | 16.87 | 13.04 | 4.87 | 23.84 | 21.39 | 20.25 | 17.41 | 17.10 |
| CHF | 49.63 | 3.86 | 6.63 | 15.26 | 26.03 | 31.02 | 17.21 | 21.53 | 21.13 | 21.07 | 18.62 | 17.65 |
| COAGULOPATHY | 46.17 | 25.16 | 15.21 | 18.97 | 19.95 | 15.69 | 5.02 | 20.97 | 20.16 | 20.34 | 18.12 | 20.41 |
| DEFANAEMIA | 31.12 | 23.38 | 13.43 | 17.22 | 20.38 | 17.89 | 7.70 | 22.67 | 20.86 | 20.36 | 18.03 | 18.08 |
| DEPRESSION | 37.70 | 44.07 | 19.15 | 15.79 | 10.49 | 7.59 | 2.90 | 25.16 | 21.36 | 19.53 | 16.62 | 17.33 |
| DIABETES | 51.38 | 19.40 | 17.74 | 23.32 | 21.09 | 14.02 | 4.43 | 23.09 | 21.20 | 20.67 | 17.44 | 17.60 |
| DRUG | 61.65 | 69.01 | 12.74 | 8.68 | 5.26 | 3.25 | 1.07 | 37.71 | 23.20 | 17.56 | 12.01 | 9.53 |
| FED | 41.23 | 9.47 | 10.65 | 19.05 | 26.27 | 25.56 | 9.00 | 22.20 | 21.21 | 20.00 | 18.42 | 18.19 |
| HYPERTENSION | 47.89 | 15.24 | 17.20 | 23.43 | 21.25 | 16.64 | 6.23 | 20.89 | 20.70 | 20.70 | 18.63 | 19.09 |
| HYPOTHYROIDISM | 20.13 | 21.25 | 17.28 | 20.97 | 18.96 | 15.25 | 6.29 | 20.85 | 21.01 | 21.04 | 18.54 | 18.56 |
| LIVER | 53.41 | 29.99 | 22.84 | 22.56 | 15.26 | 7.77 | 1.59 | 24.77 | 20.87 | 19.57 | 17.21 | 17.57 |
| LYMPHOMA | 53.88 | 11.53 | 14.21 | 24.70 | 25.59 | 18.62 | 5.34 | 18.50 | 19.08 | 20.81 | 20.42 | 21.19 |
| OBESITY | 43.70 | 35.64 | 21.07 | 21.01 | 14.26 | 6.65 | 1.37 | 24.80 | 21.73 | 19.65 | 16.98 | 16.83 |
| NEURO | 47.25 | 13.15 | 10.92 | 17.80 | 23.79 | 23.54 | 10.80 | 21.39 | 20.25 | 20.67 | 18.28 | 19.40 |
| PARALYSIS | 49.88 | 14.87 | 11.35 | 16.19 | 22.14 | 24.37 | 11.08 | 23.59 | 21.46 | 20.48 | 17.88 | 16.59 |
| PUD | 57.96 | 13.56 | 15.38 | 21.72 | 23.29 | 19.10 | 6.95 | 23.97 | 21.50 | 20.25 | 17.41 | 16.87 |
| PVD | 60.65 | 6.14 | 11.60 | 21.72 | 27.64 | 24.49 | 8.41 | 23.15 | 21.15 | 20.63 | 18.00 | 17.08 |
| PSYCHOSES | 47.01 | 37.78 | 16.24 | 15.66 | 13.62 | 11.76 | 4.94 | 27.67 | 21.45 | 19.54 | 16.24 | 15.09 |
| PCD | 46.02 | 12.46 | 13.08 | 23.01 | 25.94 | 19.47 | 6.04 | 22.36 | 20.98 | 21.15 | 18.41 | 17.09 |
| RENAL | 45.93 | 6.35 | 9.28 | 20.34 | 29.10 | 26.70 | 8.24 | 21.37 | 20.92 | 21.38 | 19.04 | 17.30 |
| CTD | 33.92 | 16.50 | 16.12 | 23.10 | 22.15 | 16.41 | 5.72 | 20.06 | 19.14 | 21.53 | 19.22 | 20.05 |
| CANCER | 48.17 | 10.02 | 14.11 | 24.38 | 24.89 | 19.48 | 7.12 | 18.71 | 19.56 | 20.77 | 19.61 | 21.34 |
| VALVULAR | 45.95 | 12.05 | 10.36 | 19.79 | 27.59 | 22.97 | 7.24 | 19.06 | 19.80 | 20.74 | 19.93 | 20.46 |
| WEIGHT | 45.16 | 20.34 | 14.20 | 19.87 | 22.62 | 17.79 | 5.18 | 25.02 | 21.17 | 19.66 | 16.90 | 17.25 |

### 10.5 Directed hypergraph weights

**Figure S5:**
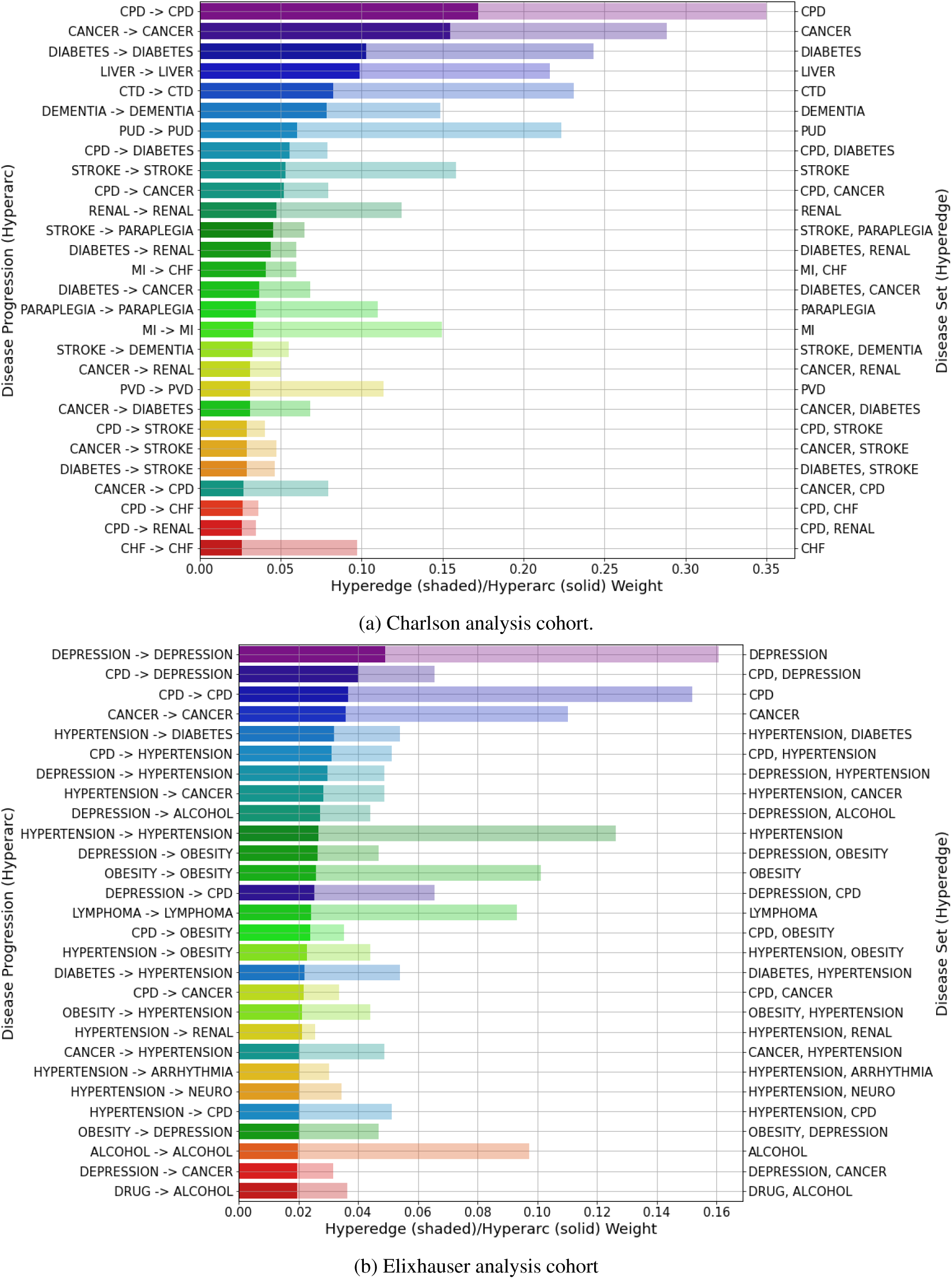
Top 28 hyperarc weights (solid) superimposed onto their parent hyperedge weight (shaded), with their titles on the left and right *y*-axes, respectively, in the Charlson (a) and Elixhauser (b) analysis cohorts. Self-transition hyperarcs represent individuals observed with only that condition.

### 10.6 Directed graph weights

**Figure S6:**
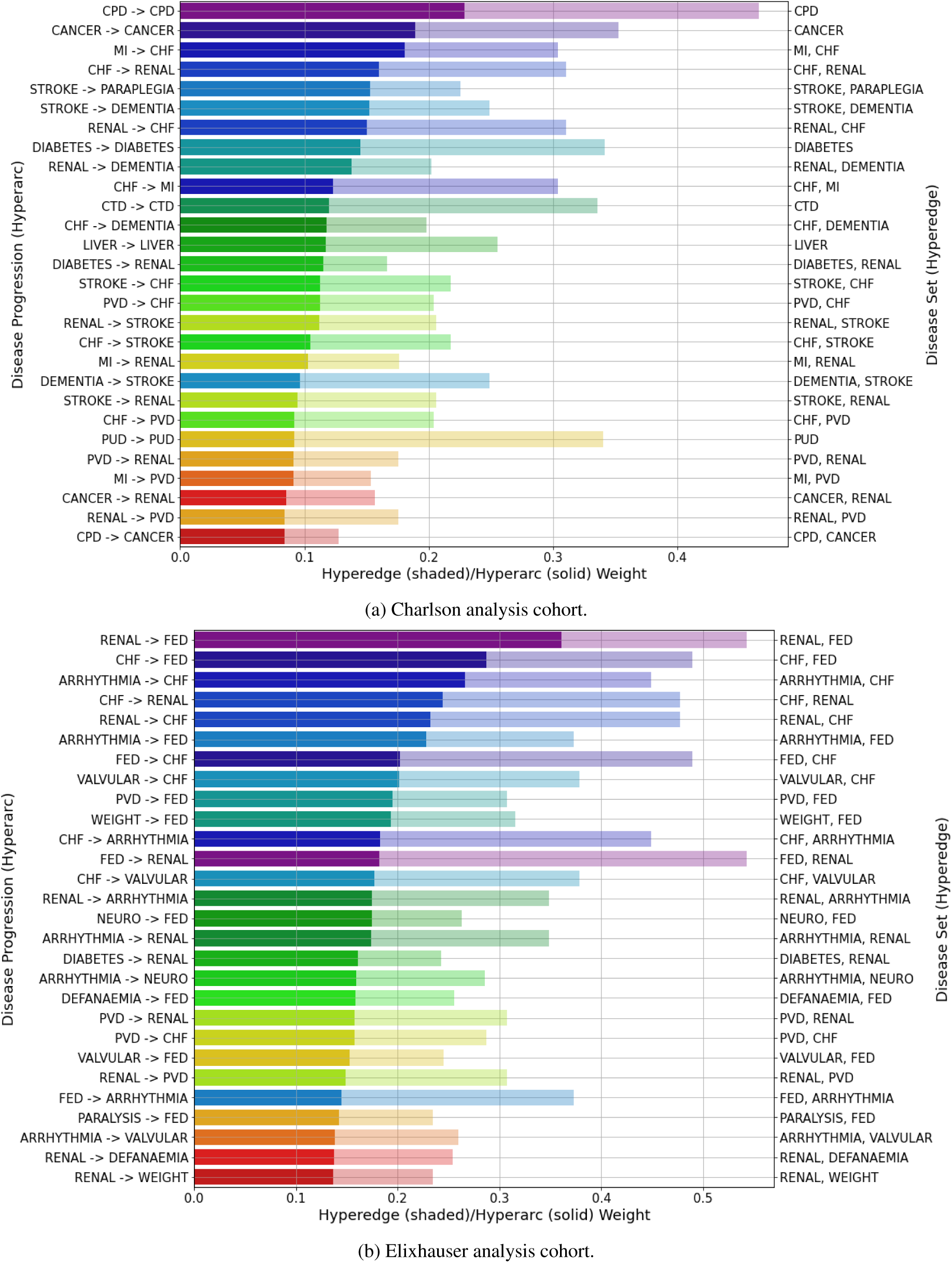
Top 28 hyperarc (solid) and hyperedge (shaded) weights for the Charlson (a) and Elixhauser (b) analysis cohorts using the directed simple graph.

### 10.7 Population-level PageRank signed differences

**Figure S7:**
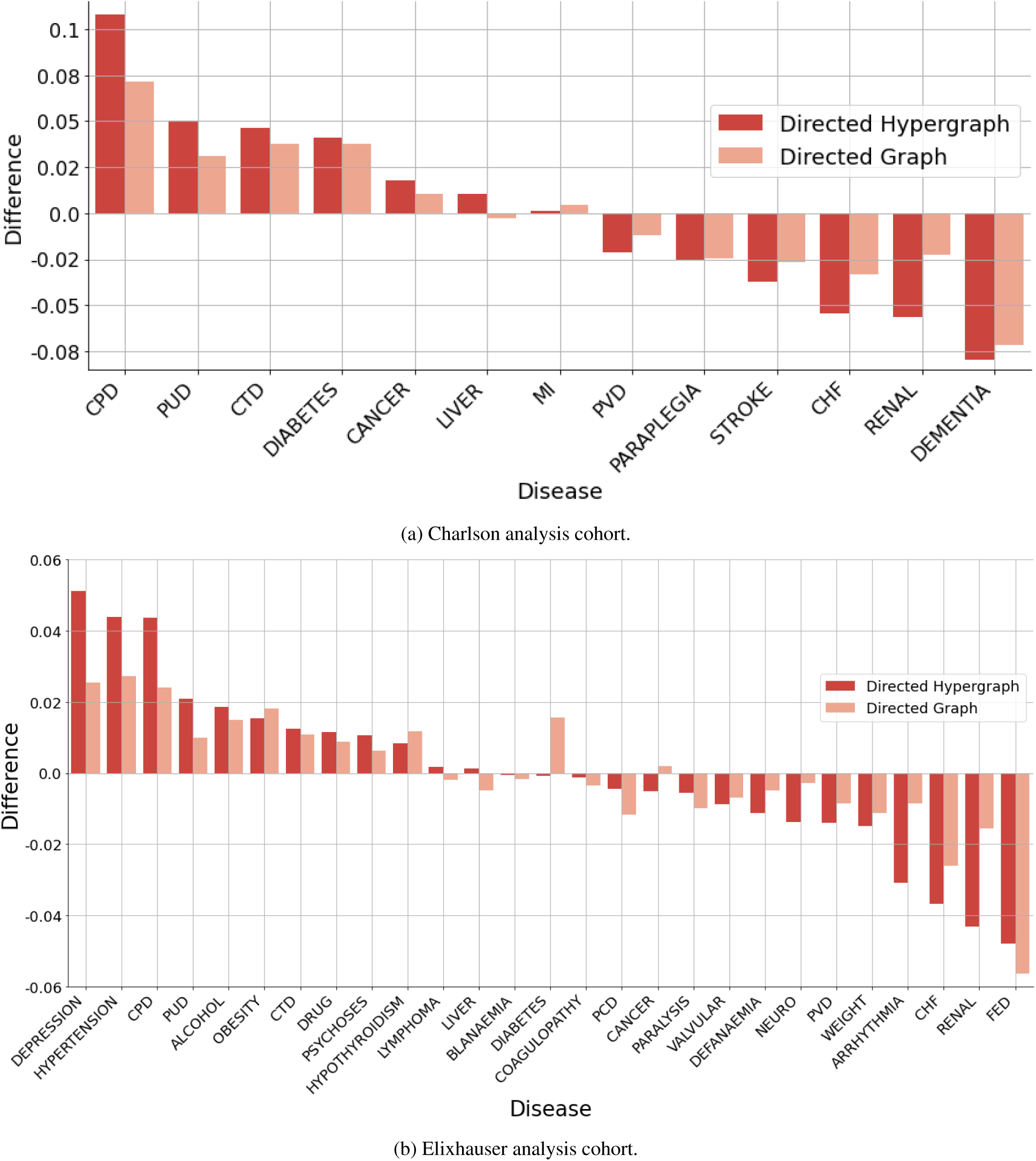
Signed difference of successor and predecessor PageRank centrality values for each disease in both analysis cohorts. Positive values represent predecessor inclined conditions and vice versa for successor conditions. Purple bars show PageRank signed differences from using the directed hypergraph and orange bars represent the directed simple graph.

### 10.8 Age-sex stratification PageRank centrality

**Figure S8:**
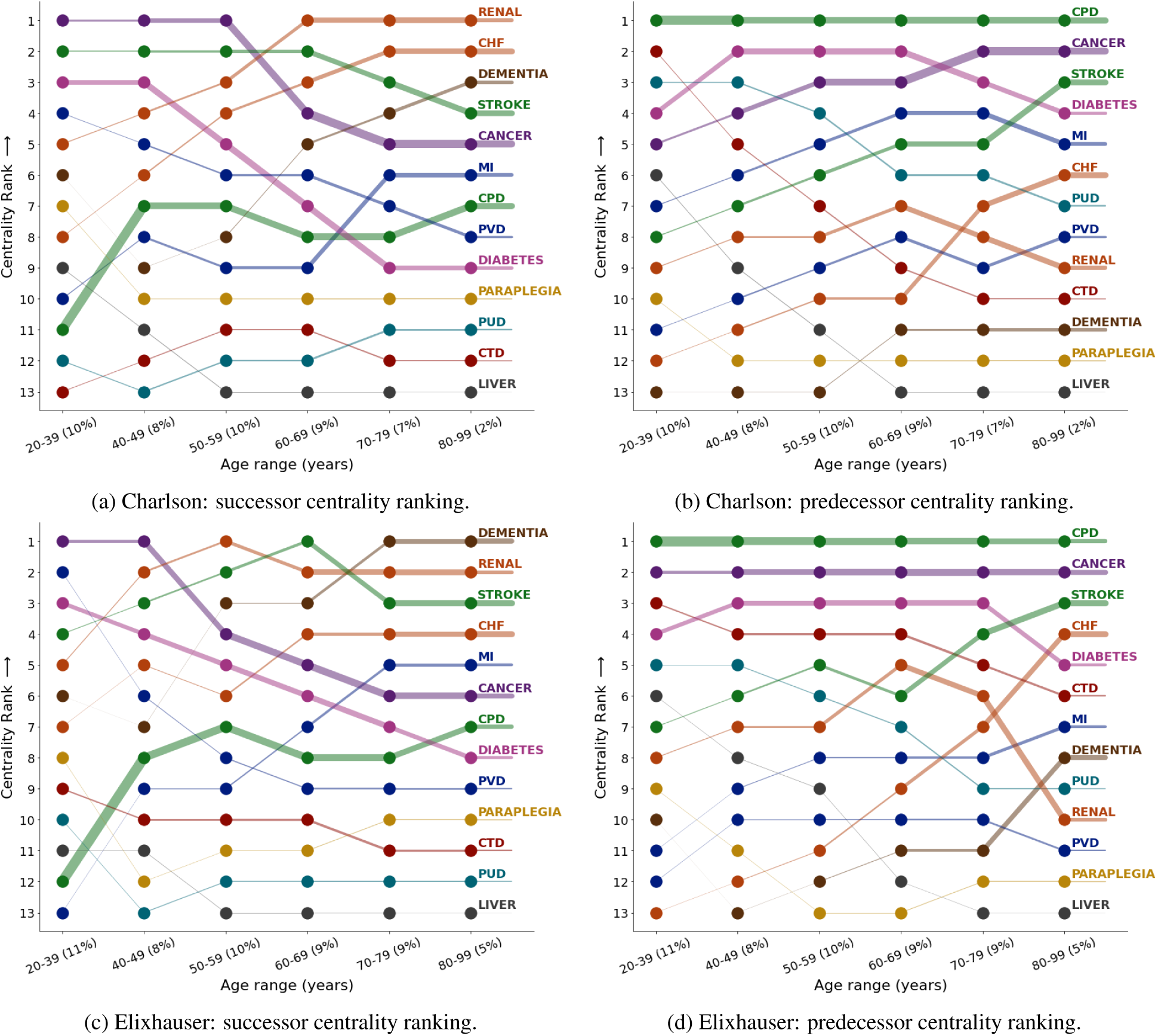
PageRank centrality ranking of diseases in the male (a–b) and female (c–d) stratification’s of the Charlson analysis cohort, split by age group.

**Figure S9:**
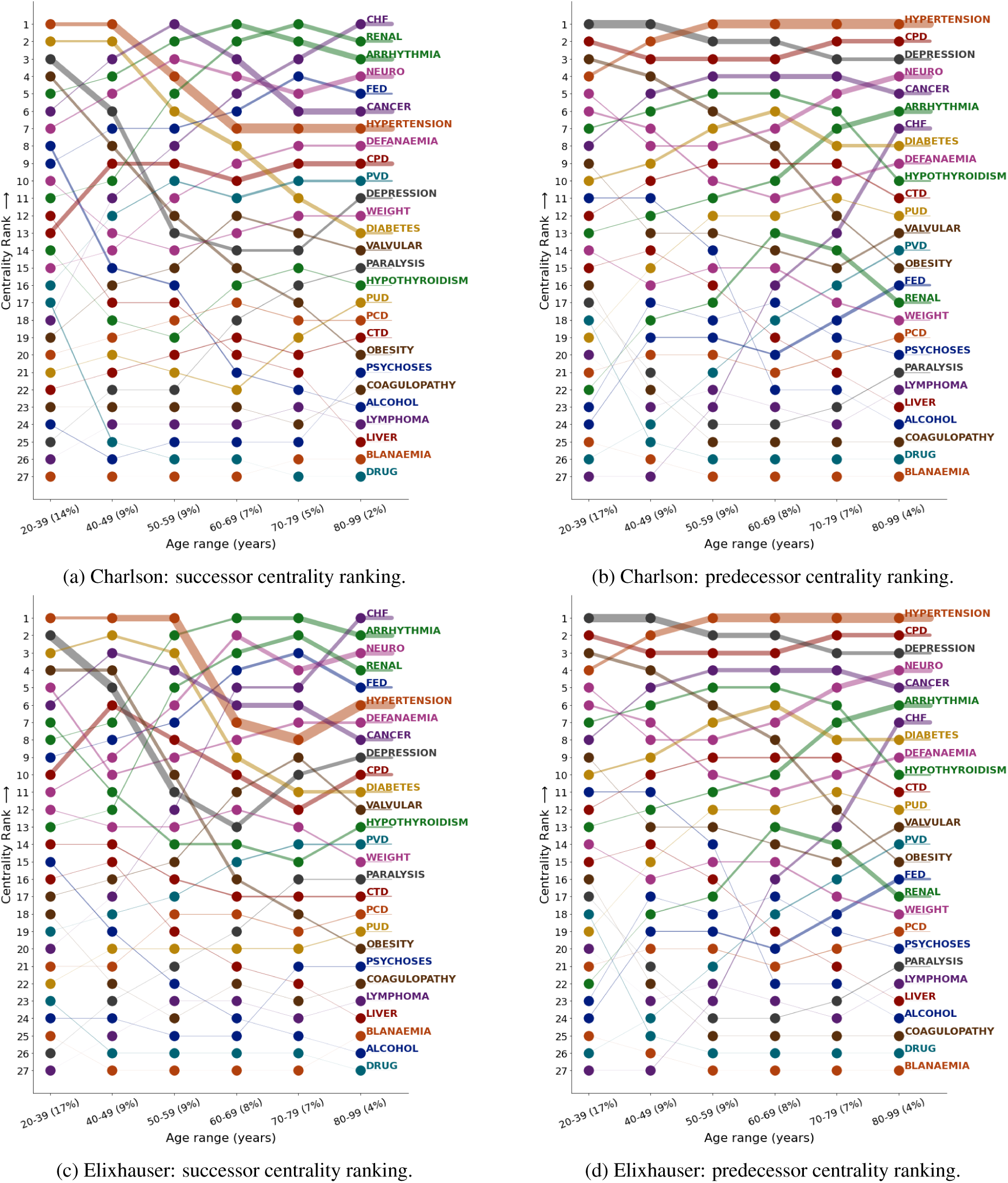
PageRank centrality ranking of diseases in the male (a–b) and female (c–d) stratification’s of the Elixhauser analysis cohort, split by age group.

## Footnotes

1 Note that the parent-child terminology used here is separate from that found in probabilistic graphical models, where parents of children nodes refer to directed, outgoing connections from parent nodes to children nodes within the same graph.

2 The formulation of the original S⊘rensen-Dice coefficient for the two-case setting can be recovered when there are only two diseases in the dataset and setting weight coefficients *w_j_* = *w_k_* = 0.5.

3 See Tran, et al. [39] for a description of the incidence, node and edge degree matrices for the directed hypergraph.

